# Epi-Clock: A sensitive platform to help understand pathogenic disease outbreaks and facilitate the response to future outbreaks of concern

**DOI:** 10.1101/2022.09.14.22279955

**Authors:** Cong Ji, Junbin (Jack) Shao

**Affiliations:** Liferiver Science and Technology Institute, Shanghai ZJ Bio-Tech Co., Ltd., Shanghai, China

**Keywords:** Divergence, Host bias, Mutation type, Codon usage bias, ZHU algorithm

## Abstract

The lack of virus fossilization precludes any references or ancestors for inferring evolutionary processes, and viruses have no cell structure, metabolism, or space to reproduce outside host cells. Most mutations yielding high pathogenicity become removed from the population, but adaptive mutations could be epidemically transmitted and fixed in the population. Therefore, determining how viruses originated, how they diverged and how an infectious disease was transmitted are serious challenges. To predict potential epidemic outbreaks, we tested our strategy, Epi-Clock, which applies the ZHU algorithm on different SARS-CoV-2 datasets before outbreaks to search for real significant mutational accumulation patterns correlated with the outbreak events. We imagine that specific amino acid substitutions are triggers for outbreaks. Surprisingly, some inter-species genetic distances of *Coronaviridae* were shorter than the intra-species distances, which may represent the intermediate states of different species or subspecies in the evolutionary history of *Coronaviridae*. The insertions and deletions of whole genome sequences between different hosts were separately associated with new functions or turning points, clearly indicating their important roles in the host transmission and shifts of *Coronaviridae*. Furthermore, we believe that non-nucleosomal DNA may play dominant roles in the divergence of different lineages of SARS-CoV-2 in different regions of the world because of the lack of nucleosome protection. We suggest that strong selective variation among different lineages of SARS-CoV-2 is required to produce strong codon usage bias, significantly appear in B.1.640.2 and B.1.617.2 (Delta). Interestingly, we found that an increasing number of other types of substitutions, such as those resulting from the hitchhiking effect, have accumulated, especially in the pre-breakout phase, even though some previous substitutions were replaced by other dominant genotypes. From most validations, we could accurately predict the potential pre-phase of outbreaks with a median interval of 5 days before. Using our pipeline, users may review updated information on the website https://bioinfo.liferiver.com.cn with easy registration.

## Introduction

Determining how viruses originated and diverged and how a disease was transmitted are extremely important for understanding viral disease outbreaks and facilitating responses to future outbreaks. Only 13.7% of the areas on Earth are suitable for human use, and the remaining areas are enriched with over millions of species of viruses, with an extremely large population scale and accumulations of genetic mutations, especially in the ocean. Viruses are the communication bridges between humans and nature, regardless of whether the effect is positive, so each communication process should be co-evolutionary. Since viruses lack fossilization, preventing the availability of any references or ancestors for inferring evolutionary processes, some theories about the origin of viruses have been reported, such as the degeneracy theory, DNA escape from plasmids or transposons, and viroid or satellite viruses[1]. The origin of new genes with novel functions creates genetic and phenotypic diversity in organisms, which gradually acquire increasing numbers of gene partners[2]. A non-parametric approach for estimating the date of origin of genetic variants in large-scale sequencing datasets has been developed, with allele age estimates providing a rapid approach for inferring the ancestry shared between individual genomes, determining genealogical relationships at different points in the past, and describing and exploring the evolutionary history of modern human populations[3]. However, viruses, agents on the edge of life, have the same genes as living organisms and undergo evolution by natural selection and reproduction by self-assembly but have no cell structure, metabolism, or space to reproduce except in the host cells. To study the macroevolution of viruses, several methods have been proposed, i.e., phylogenetic[4], neutral selection[5], and divergence[6] approaches. Most high-pathogenicity mutations go extinct in the population, but adaptive mutations can be fixed in the population, such that most intra-species mutations should be under neutral selection[5]. To answer the above crucial questions about outbreaks, sequencing of viral samples and supplementation of epidemiological methods could play an important role in providing nucleotide-level resolution data for outbreak-causing pathogens[7].

How does viral evolution occur across hosts? Most emerging infectious diseases in humans originate from animal reservoirs, and it is necessary to understand how and why some pathogens become capable of crossing host species barriers. Population genetic analyses provide estimates suggesting that the putative introduced genetic sequence within the RBD is evolving under directional selection, which may result in the adaptation of the virus to hosts. Unsurprisingly, the putative recombination region in the S protein was found to be highly diverse among strains from bats[8]. Emerging viral diseases are often the product of a host shift, where a pathogen jumps from its original host into a novel species. Host shifts are frequent in the evolution of most pathogens, but why pathogens successfully jump between some host species but not others is only just becoming clear[9]. Interestingly, the same sequence changes are often seen each time a virus infects a particular host. These changes may come at a cost to other aspects of the pathogen’s fitness, and this may sometimes prevent host shifts from occurring[9]. Therefore, evolution could help viruses overcome species barriers, principally by affecting virus‒host interactions; however, evolving the capability for sustained transmission in a new host species represents a major adaptive challenge because the number of mutations required is often large[10]. Truly understanding disease emergence requires a new mechanistic and integrated view of the factors that allow or prevent viruses from spreading in novel hosts, suggesting that both ecological and genetic aspects of virus emergence could be placed within a simple population genetic framework[11]. The continuous struggle between viruses and their hosts to maintain at least a constant fitness level leads to the development of an unceasing arms race, where weapons are often shuttled between the participants[12]. Variables including genome length, genome type, and recombination frequency have little predictive power[13]. The emergence of these and many other human diseases occur when an established animal virus switches to human hosts and is subsequently transmitted within human populations, while transfers between different animal hosts lead to the analogous emergence of epizootic diseases. The importance of viral host switching was underscored by the recent avian epizootics of high-pathogenicity strains of H5N1 influenza A, in which hundreds of spillover human cases and deaths have been documented[14]. The movement of EBOV within the region and viral evolution during prolonged human-to-human transmission have been reported[15]. The specific amino acid substitutions in the EBOV GP have increased tropism for human cells while reducing tropism for bat cells. Such increased infectivity may have enhanced the ability of EBOV to transmit among humans and contributed to the wide geographic distribution of some viral lineages[16]. Hence, it seems reasonable to conclude that if there is a positive relationship between mutations in viruses and the possibility of adaptation to the host, the association between the divergence of viruses and adaptation allowing host transfer is under strong selection.

Remarkably, it has been reported that nucleosomes suppress spontaneous mutations in a base-specific manner in eukaryotes such that nucleosome occupancy nearly eliminated cytosine deamination, resulting in an ∼50% decrease in the C->T mutation rate in nucleosomal DNA[17]. However, the organization of genomic DNA into defined nucleosomes, which has long been viewed as a hallmark of eukaryotes, has been challenged by the identification of “minimalist” histones in archaea and, more recently, by the discovery of genes that encode fused remote homologues of the four eukaryotic histones in *Marseilleviridae*, a subfamily of giant viruses that infect amoebae. That is, viral doublet histones are essential for viral infectivity, localize to cytoplasmic viral factories after virus infection and ultimately are found in mature virions[18]. It has also been reported that giant viruses belonging to the nucleocytoplasmic large DNA virus (NCLDV) group have histone-like genes in their genomes. One possibility is histone doublets in MVs, and a nucleosomal structure may have been acquired to function in the infected cell for protection of the viral genome from detection and degradation and/or as a means of interfering with host cell regulation during infection, which seems reasonable given that host‒virus arms races are a powerful source of adaptation, and most genes gained by NCLDVs from their different hosts are likely linked to viral defences[19].

At present, the phylogenetic network faithfully traces routes of infections for documented coronavirus disease 2019 cases, indicating that such networks could likewise be successfully used to help trace undocumented SARS-CoV-2 infection sources, which could then be quarantined to prevent recurrent spread of the disease worldwide[20]. To compare virus genomes before epidemic and after epidemics, it is necessary to search for the accumulation pattern of pathogenic mutations contributing to the outbreak. Recently, it has been reported that the Global Epidemic and Mobility Model can provide an estimate of seasonal transmission potential by running a Monte Carlo maximum likelihood analysis to generate stochastic simulations of epidemic spread worldwide[21]. Epi-Factors, a manually curated database, provides information about epigenetic regulators and their complexes, targets and products[22]. Some of these mutations have presumably arisen as a result of the virus evolving under immune system selection pressure in infected individuals, and at least one, lineage B.1.1.7, potentially resulted from a chronic infection[23]. It has been reported that SARS-CoV-2 Lambda, a new variant of interest, is now spreading in some South American countries. The spike protein of the Lambda variant is more infectious and was attributed to the T76I and L452Q mutations[24]. It has been recently reported that there have been multiple waves of replacement between strains, with the first wave being a group of four mutations (C241T, C3037T, C14408T and A23403G). This DG (D614G) group, which was fixed at the start of the pandemic, is the foundation of all subsequent waves of strains. The European DG1111strain had acquired the highly adaptive DG mutations in pre-pandemic Europe and had been spreading in parallel with the Asian strains[25]. Because emerging pathogens have the potential to impose substantial mortality, morbidity and economic burdens on human populations, tracking the spread of infectious disease to assist in their control has traditionally relied on the analysis of case data gathered as an outbreak proceeds[26]. Based on the requirements and challenges, including the need for flexible platforms that generate sequence data in real time and for these data to be shared as widely and openly as possible[26], here, we demonstrate our strategy, with the aim of developing a sensitive process for predicting the potential trigger of outbreaks to facilitate the response to future outbreaks of concern.

## Methods

To explore divergence of the whole family of *Coronaviridae*, we computed intra-species and inter-species p-distances of whole genome sequences by MEGA11 and plotted the distances by the boxplot function of R. To illustrate the atlas of new ages within different populations, we applied a genome alignment-based pipeline to infer the origin time of a given genomic region by a 6 bp sliding window with numbers of dated variants, including insertions in red and deletions in blue. We scanned the whole genome sequences with 6 bp sliding windows and summarized the mean values of mutation rates of different mutation types. According to the reference genomes among different hosts, we demonstrated host shifts within 6 bp sliding windows in the evolution of *Coronaviridae* and presented them by Circos.

Similarly, we demonstrated nucleotide mutations and amino acid substitutions among different SARS-CoV-2 lineages. We computed mutation rates by sliding windows for different mutation types. Then, we presented the 144 amino acid substitution distributions of different lineages by GeneWise. The codon usage numbers were converted into relative synonymous-codon usage (RSCU) values[28], which was simply the observed frequency of a codon divided by the frequency expected under the assumption of equal usage of the synonymous codons for an amino acid[28] [27].

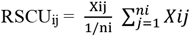

A neighbour-joining phylogenetic tree of separate lineages of SARS-CoV-2 was constructed by MEGA v11.0.11. To predict the potential epidemic mutation patterns in the severely affected areas, we summarized testing data from OWID (Our World in Data) until Feb 7th 2022 that were continually collated from official government sources worldwide. We plotted the frequency distribution of confirmed cases of SARS-CoV-2 in different severely affected areas, such as Africa, Asia, Europe, North America, Oceania, and South America, along the timeline. At the same time, we set the baseline of new cases per millions of substitutions as the internal control group and the widespread types of substitutions as the external control group so that we could exclude other effects on epidemic outbreak events. We excluded areas without any outbreak time, as labelled by the red box, and searched the mutation patterns appearing to be related to the outbreak until the appearance of the Omicron variant because this corresponded with the time of universal immunization. To ensure the true values of the outbreak of new cases of SARS-CoV-2, we carefully detected peaks following strict standards; that is, the values of new cases per million had to be above 30. Then, we separately split the sets of population samples from 1 day to 30 days before outbreaks based on the same location according to 144 amino acid substitutions or deletions shown in Figure 2b and Supplementary Table 2. The dataset contained 13740300 observations, including 6300 true sets and 2181 features, which amounted to 117 different countries or regions on all five continents, divided into the 75 training sets and 42 validation sets listed in Supplementary Table 3. Then, we performed GLM (generalized linear model) analysis on individual countries/regions to find the optional mutational patterns as in the training phase. Across N generations of training by GLM and reordering, we found 171 significant substitutions as potential epi-factors within 55 different countries and regions. Finally, we assessed the ZHU prediction by 42 validation sets with precision, recall, and accuracy, as follows.

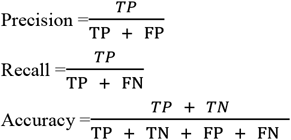

## Results

### Divergence of the whole *Coronaviridae* family within different populations and adaptation to cross host species barriers

To comprehensively explore the evolution of the whole family of *Coronaviridae*, we performed a genomic comparison of *Coronaviridae* to search for new age patterns related to virulence or transmissibility, as shown in Supplementary Figure 1. In the analysis of intra-species and inter-species genetic distances of whole genome sequences of *Coronaviridae*, it was obvious that the intra-species genetic distances were lower, as shown in Supplementary Figure 1. For instance, the sequencing similarities of SARS-CoV-2 and bat-SL-CovZC45 and SARS-CoV-2 and bat-SL-CovZXC21 are nearly 88%; however, the similarities of SARS-CoV-2 and SARS-CoV and SARS-CoV-2 and MERS are 79% and 50%, respectively. According to a comparison of the genetic distances of H1N1 and H3N2 with those of SARS-CoV-2 and SARS-CoV, we found that the p-distance of SARS-CoV-2 and Bat SARS-like CoV was nearly 0.13 and the p-distance of SARS-CoV-2 and SARS-CoV was 0.24 in MEGA[28]. Both of these values are lower than the p-distances of H1N1 and H3N2 (0.8) and H1N1 and H7N9 (0.73). As shown in Supplementary Figure 1, it was obvious that most inter-species genetic distances of *Coronaviridae* are longer than intra-species distances, i.e., SARS-CoV-2, SARS-CoV, SADS, NL63, MERS, London1, HKU5, HKU4, HKU3, HKU2, and BATS.

Conversely, some inter-species genetic distances of *Coronaviridae* are shorter than the intra-species genetic distances, such as those for OC43 and 229E, which may be the intermediate states of different species or subspecies in the whole evolutionary history of *Coronaviridae*. It suggested that these probably represent another independent acquisition though either specific horizontal gene transfer or recombination events[29-31]. The relationship between variants and frequency in different geographical regions has been characterized, and the value of using age information in interpreting variants of functional and selective importance, such as using allele age estimates to power a rapid approach for inferring the ancestry shared between individual genomes in order to quantify genealogical relationships, has been demonstrated[3]. Here, we explore the divergence time of species or sub-species of *Coronaviridae*, which presented the diversity of such ages within populations separately associated with new functions or important turning points. Interestingly, we explored the dated variants of different species or subspecies and generated an atlas of new ages with SARS-CoV-2 as the reference genome in Supplementary Figure 2. It was demonstrated that SARS-CoV, HKU3, and BATS are very close to SARS-CoV-2 and far away from OC43 and HKU1 on the S protein, although there are 5 kb insertions of BATS in Orf7a, Orf7b, and Orf8 and 15 kb deletions of OC43, HKU9, and HKU1 in Orf7a, Orf7b, Orf8 and N. All populations show evidence of insertions, deletions or substitutions that have driven the divergence of the whole family of *Coronaviridae* in response to natural selection, random genetic drift, host gene editing, viral proofreading[32].

Most strikingly, as shown in Figure 1 and Supplementary Table 1, the insertions and deletions of whole genome sequences between different hosts should play an important role in host transmission and the shift of *Coronaviridae* presented by Circos. For instance, human coronavirus 229E in *Camelus* and *Vicugna pacos* have S protein sequences similar to those observed in *Homo sapiens*, while having 500 bp deletions in *Hipposideros* and *Macronycteris vittata*. When used as the reference genome, *Rhinacovirus* in *Sus scrofa* was similar to that in *Rhinolophus ferrumequinum* and divergent from that in *Rhinolophus affinis* in that insertions and deletions all appear in the S protein, which may be related to host bias. It is likely that accumulations of insertions and deletions of alphacoronavirus, human coronavirus OC43, Middle East respiratory syndrome-related coronavirus, severe acute respiratory syndrome-related coronavirus, *Deltacoronavirus*, and avian coronavirus emerged on the S protein. The similarity of SARS-CoV-2 S proteins in *Rhinolophus sinicus, Rhinolophus macrotis, Canis lupus familiaris*, and *Panthera tigris jacksoni* is very high with the SARS-CoV-2 S protein in *Homo sapiens*, which means that the ancestral hosts of SARS-CoV-2 in *Homo sapiens* should be closely related to the species as detailed in Supplementary Figures 3-10. Wildlife host species richness has been proposed as an important predictor of disease emergence. Similarly, host populations with low biodiversity might harbour fewer viruses and an increased risk of emergence. Conversely, high host biodiversity has also been linked to a decrease in disease risk through the ‘dilution effect’[11]. Advances in deep sequencing have made it possible to measure within-host genetic diversity in both acute and chronic influenza infections[33]. Factors such as antigenic selection, antiviral treatment, tissue specificity, spatial structure, and multiplicity of infection may affect how influenza viruses evolve within human hosts[33]. In summary, the more mutations viruses have, the greater the possibility of adaptation allowing them to cross host species barriers, which indicates that increased host diversity will protect against the emergence of pathogens.

**Fig. 1.**
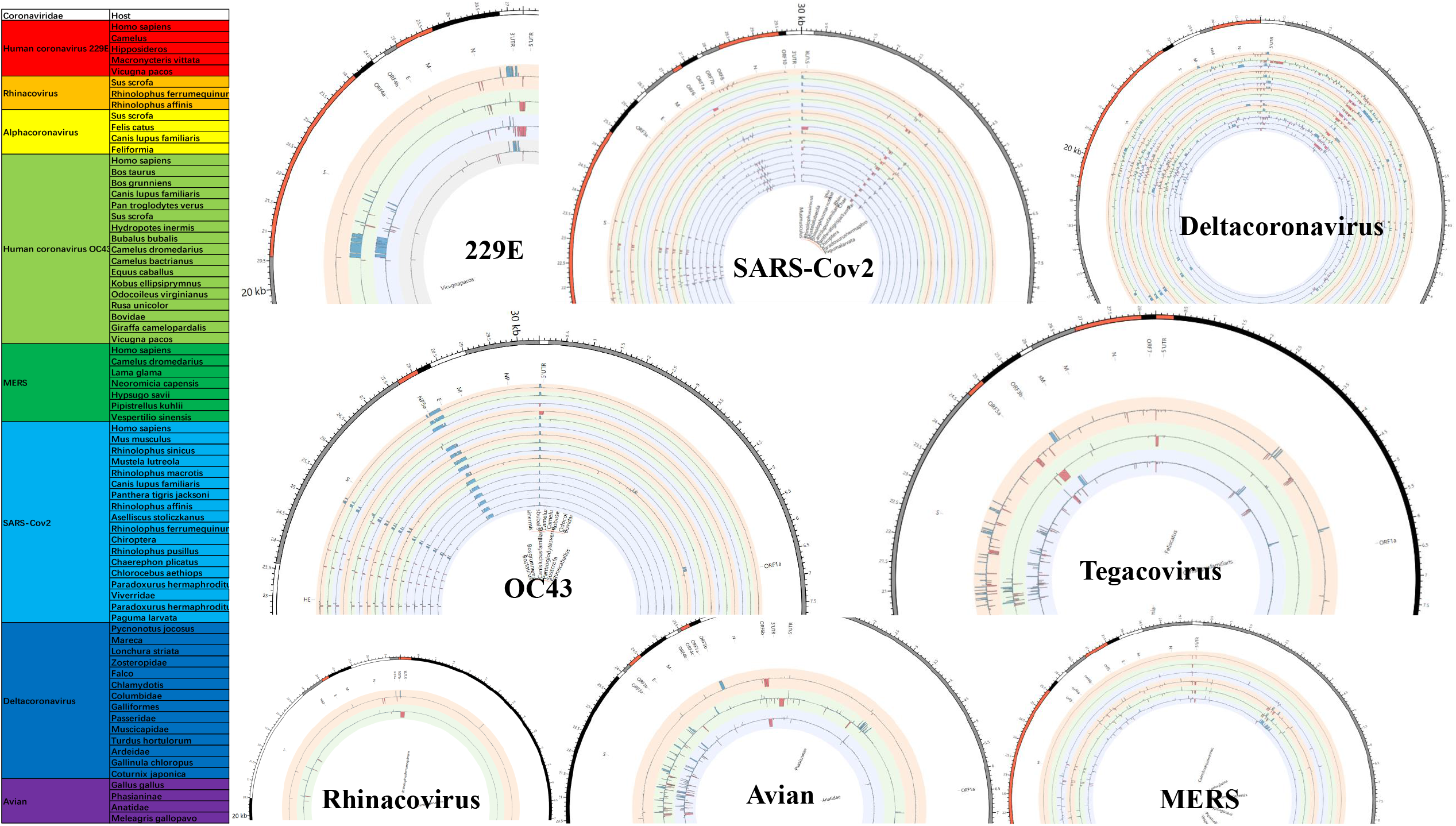
Host shifts in the evolution of *Coronaviridae*. Here, we partly present the divergence of whole genome among different hosts in different species or subspecies of *Coronaviridae*, such as human coronavirus 229E in *Camelus, Hipposideros, Macronycterisvittata, Vicugnapacos* compared with in *Homo sapiens*; SARS-CoV-2 in *Mus musculus, Rhinolophus sinicus, Mustela lutreola, Rhinolophus macrotis, Canis lupus familiaris, Panthera tigris jacksoni, Rhinolophus affinis, Aselliscus stoliczkanus, Rhinolophus ferrumequinum, Chiroptera, Rhinolophus pusillus, Chaerephon plicatus, Chlorocebus aethiops, Paradoxurus hermaphroditus, Viverridae, Paradoxurus hermaphroditus, Paguma larvata* compared with in *Homo sapiens*; Deltacoronavirus in *Mareca, Lonchura striata, Zosteropidae, Falco, Chlamydotis, Columbidae, Galliformes, Passeridae, Muscicapidae, Turdus hortulorum, Ardeidae, Gallinula chloropus, Coturnix japonica;* Human coronavirus OC43 in *Bos Taurus, Bos grunniens, Canis lupus familiaris, Pan troglodytes verus, Sus scrofa, Hydropotes inermis, Bubalus bubalis, Camelus dromedaries, Camelus bactrianus, Equus caballus, Kobus ellipsiprymnus, Odocoileus virginianus, Rusa unicolor, Bovidae, Giraffa Camelopardalis, Vicugna pacos;* Alphacoronavirus in *Felis catus, Canis lupus familiaris, Feliformia* compared with in *Sus scrofa*; Rhinacovirus in *Rhinolophus ferrumequinum, Rhinolophus affinis* compared with in *Sus scrofa*; Avian in *Phasianinae, Anatidae, Meleagris gallopavo* compared with in *Gallus gallus*; Middle East respiratory syndrome-related coronavirus in *Camelus dromedaries, Lama glama, Neoromicia capensis, Hypsugo savii, Pipistrellus kuhlii, Vespertilio sinensis* compared with in *Homo sapiens*.

### The distribution of rates of different mutation types and codon usage bias in different lineages of SARS-CoV-2

Pathogens have always been a major cause of human mortality, so they impose strong selection pressure on the human genome[34]. Across different lineages of SARS-CoV-2, most nucleotide substitutions were found in the regions of the S gene and N gene as presented in Supplementary Figure 11, which clearly demonstrates that accumulations of mutations drove the evolutionary history of different lineages of SARS-CoV-2. Then, we demonstrated the richness distribution of different mutation types across the whole genomes, in which mutation rates of C->T, G->T and T->C were dominant in driving the divergence of different lineages of SARS-CoV-2, as shown in Figure 2a and Supplementary Figures 12-17. In particular, the mutation rate distributions of C->T are highly enriched in all lineages of SARS-CoV-2, and the mutation rate distributions of G->T are relatively enriched in B.1.1.318, B.1.1.529 (Omicron), B.1.617.1 (Kappa), B.1.617.3, B.1.621 (Mu), B.1.640.1, B.1.640.2, C.1.2, C.37 (Lambda), and P.3 (Theta).

**Fig. 2.**
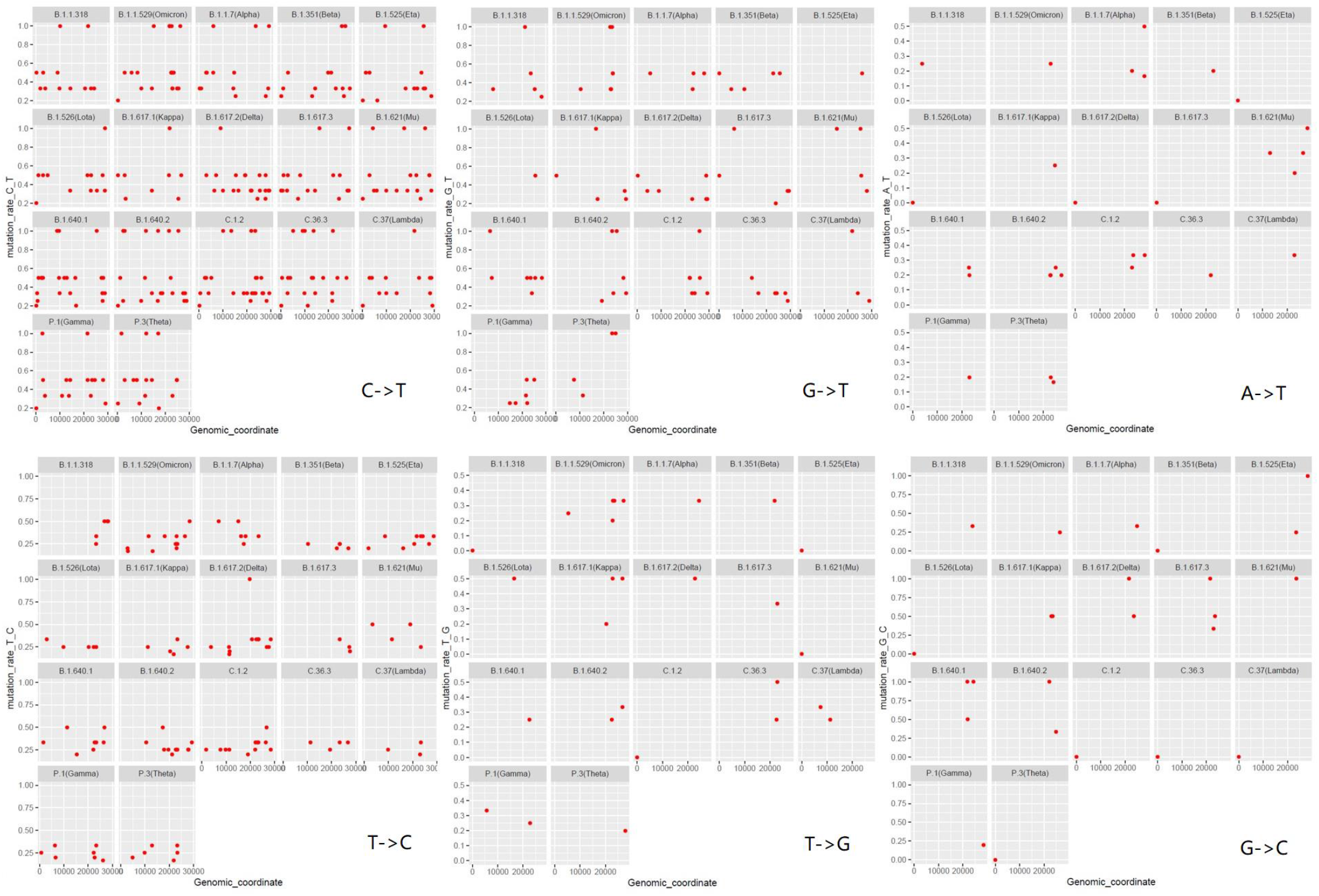
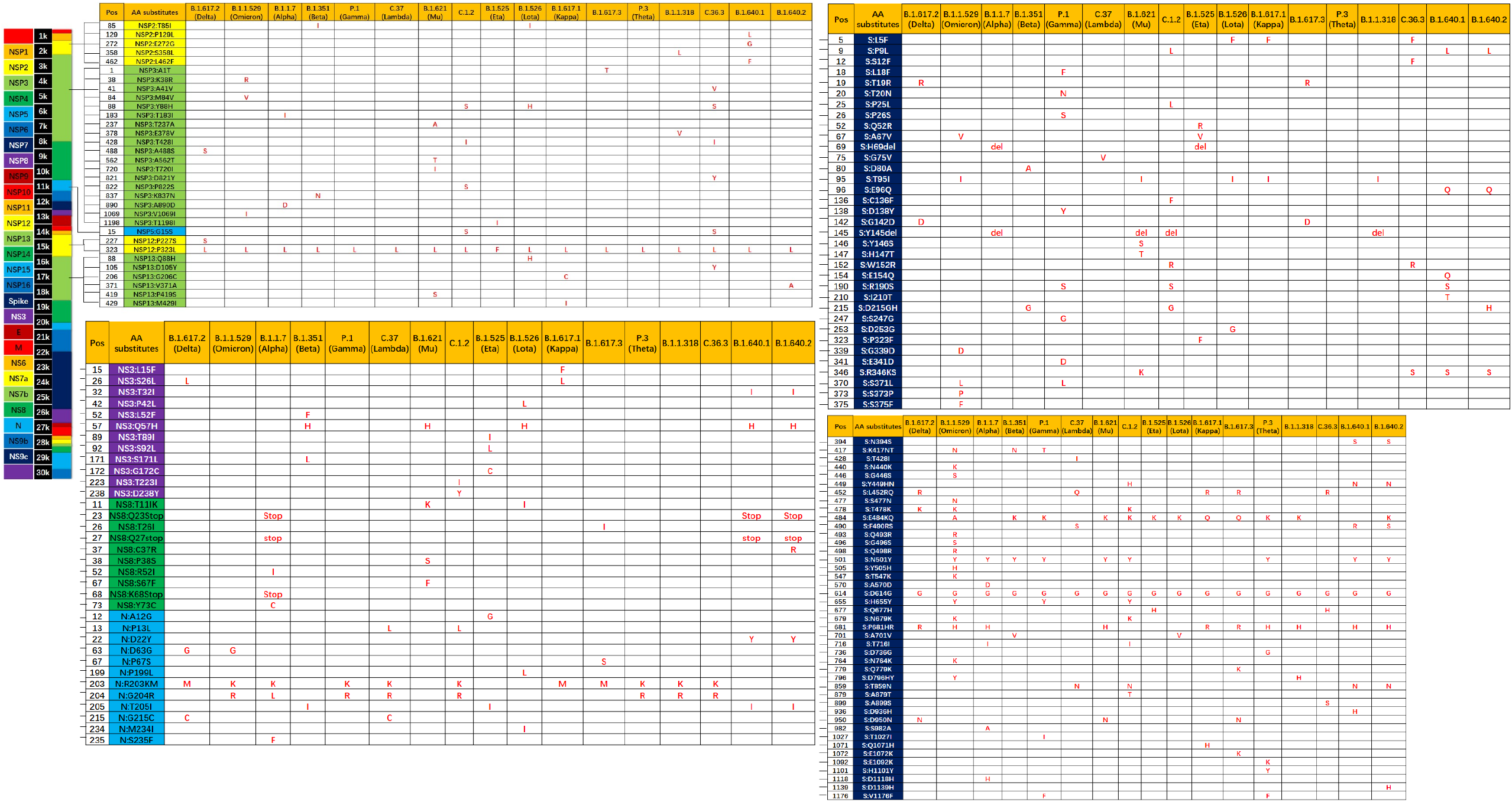
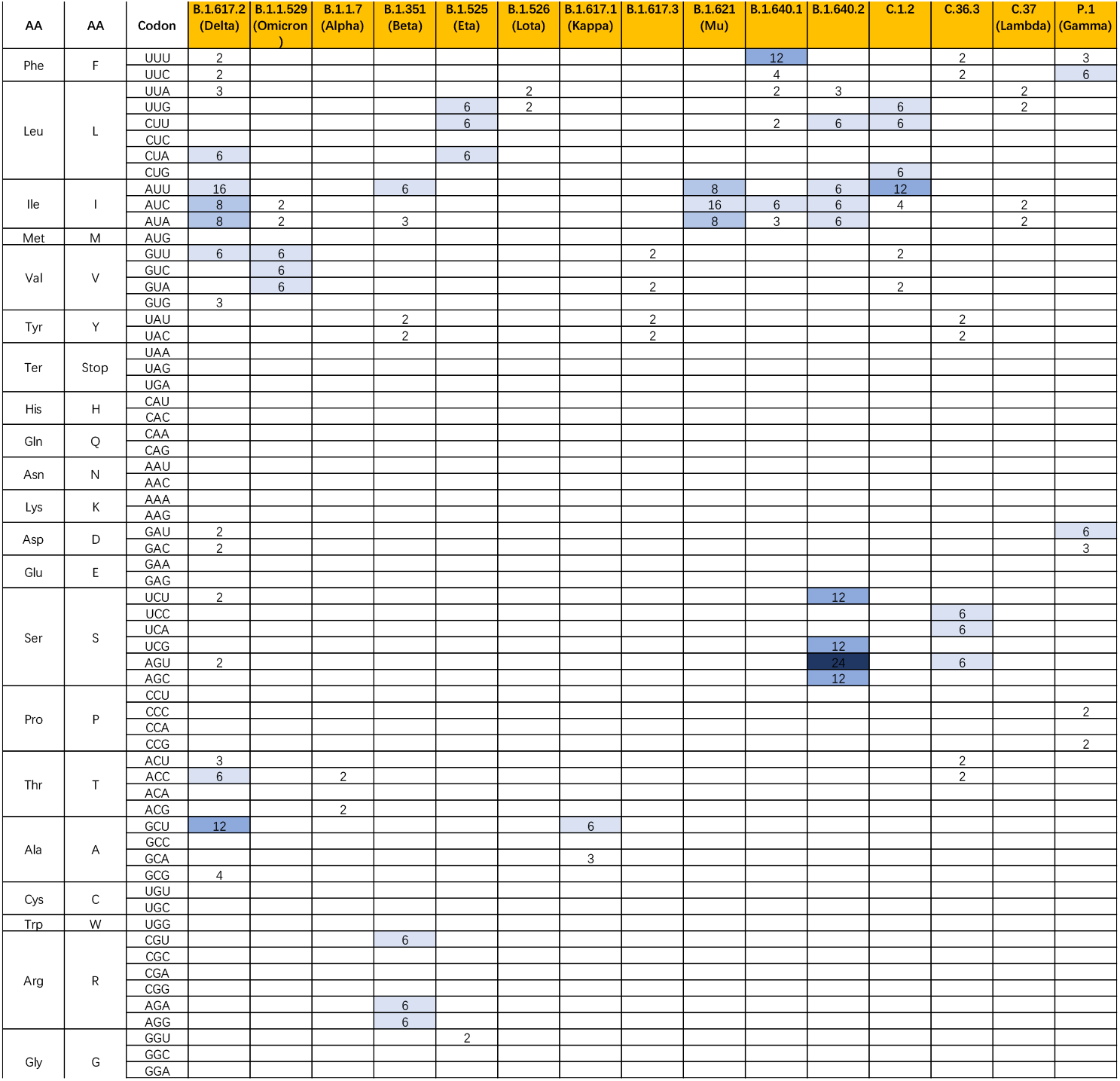
The distribution of mutation rates for different mutation types and codon usage bias of different lineages of SARS-CoV-2. **2a**. Different lineages are represented by different mutation types, i.e., C->T, G->T, A->T, T->C, T->G, G->C. **2b**. The distribution of amino acid substitutions among different SARS-CoV-2 lineages. **2c**. RSCU values in different lineages of SARS-CoV-2.

Conversely, the patterns of A->T, T->G, G->C are relatively rare, and it was clear that some points with high mutation rates in the A->T distribution derive from B.1.1.7 (Alpha) and B.1.621 (Mu); several points with high mutation rates in the T->G distribution from B.1.526 (Lota), B.1.617.1 (Kappa), B.1.617.2 (Delta), and C.36.3; and some points in the G->C distribution from B.1.525 (Eta), B.1.617.2 (Delta), B.1.617.3, B.1.621 (Mu), B.1.640.1 and B.1.640.2. Recently, some study reported that N proteins combined in vitro with short fragments of the viral genome to form 15-nm particles similar to the vRNP structures observed within virions. These vRNPs depend on regions of N protein that promote protein-RNA and protein-protein interactions. Phosphorylation of N protein in its disordered serine/arginine region weakens these interactions to generate less compact vRNPs to support other N protein functions in viral transcription[35]. Therefore, we believe that nucleosomal hereditary material (DNA/RNA) undergoes fewer C->T mutations in SARS-CoV-2. This nucleosomal hereditary material (DNA/RNA) is protected by the nucleosomal structure, with viral doublet histones essential for viral infectivity and viral defences leading to multiple host adaptation/transmission. In contrast, non-nucleosomal deoxyribonucleic acid may play dominant roles in the divergence of various lineages of SARS-CoV-2 in different regions of the world without the protection of nucleosomes.

Furthermore, we present the recent amino acid substitutions among different SARS-CoV-2 lineages, including B.1.617.2 (Delta), B.1.1.529 (Omicron), B.1.1.7 (Alpha), B.1.351 (Beta), P.1 (Gamma), C.37 (Lambda), B.1.621 (Mu), C.1.2, B.1.525 (Eta), B.1.526 (Lota), B.1.617.1 (Kappa), B.1.617.3, P.3 (Theta), B.1.1.318, C.36.3, B.1.640.1, and B.1.640.2, with a focus on NSP2, NSP3, NSP5, NSP12, NSP13, Spike, NS3, NS8, and N proteins in Figure 2b and Supplementary Table 1, which have strong effects on the divergence and evolutionary history of different lineages. This is especially true for the Spike protein, which contains a receptor-binding domain (RBD), a fusion domain and a transmembrane domain, and the NSP3 protein that is the N-terminus of coronavirus SARS-CoV non-structural protein 3 (Nsp3) and related proteins. Interestingly, compared with the dominant amino acid substitutions, i.e., NSP12:P323L and S:D614G, there seem to be unique and specific amino acid substitutions in one lineage relative to another lineage. Examples include the NSP2:P129L, NSP2:E272G, NSP2:L462F, S:E154Q, S:I210T, and S:D936H substitutions in the B.1.640.1 lineage; NSP13:V371A, S:P129L, S:D1139H, and NS8:C37R substituted in the B.1.640.2 lineage; NSP2:S358L, and NSP3:E378V substitutions in the B.1.1.318 lineage; NSP3:A41V, NSP3:D821Y, NSP13:D105Y, S:S12F, S:G212V, and S:A899S substitutions in the C.36.3 lineage; S:K2Q, S:L280F, S:G313S, S:A368V, S:D736G, S:E1092K, and S:H1101Y substitutions in the P.3 (Theta) lineage; NSP3:A1T, S:Q779K, S:E1072K, NS8:T26I, and N:P67S substitutions in the B.1.617.3 lineage; NSP13:G206C, NSP13:M429I, S:Q1071H, and NS3:L15F in the B.1.617.1 (Kappa) lineage; NSP13:Q88H, S:D253G, NS3:P42L, N:P199L, N:M234I, S:D253G, NS3:P42L, N:P199L, and N:M234I substitutions in the B.1.526 (Lota) lineage; NSP3:T1198I, S:Q52R, S:P323F, S:Q677H, NS3:T89I, NS3:S92L, NS3:G172C, and N:A12G substitutions in the B.1.525 (Eta) lineage; NSP3:P822S, S:P25L, S:C136F, S:A879T, NS3:T223I, and NS3:D238Y substitutions in the C.1.2 lineage; NSP3:T237A, NSP3:A562T, NSP3:T720I, NSP13:P419S, S:Y146S, S:H147T, S:T205I, S:R346K, NS8:P38S, and NS8:S67F substitutions in the B.1.621 (Mu) lineage; S:P13L, S:G75V, S:V76I, and S:T428I substitutions in the C.37 (Lambda) lineage; S:L18F, S:T20N, S:P26S, S:D138Y, S:S247G, S:E341D, S:S371L, S:K977Q, and S:T1027I substitutions in the P.1 (Gamma) lineage; NSP3:K837N, S:D80A, NS3:L52F, and NS3:S171L substitutions in the B.1.351 (Beta) lineage; NSP3:T183I, NSP3:A890D, S:A570D, S:S982A, S:D1118H, NS8:R52I, NS8:K68Stop, NS8:Y73C, and N:S235F substitutions in the B.1.1.7 (Alpha) lineage; NSP3:K38R, NSP3:M84V, NSP3:V1069I, S:G339D, S:S371L, S:S373P, S:S375F, S:N440K, S:G446S, S:S477N, S:Q493R, S:G496S, S:Q498R, S:Y505H, S:T547K, and S:N764K substitutions in the B.1.1.529 (Omicron) lineage; and NSP3:A488S, NSP12:P227S, and S:P77L substitutions in the B.1.617.2 (Delta) lineage.

Provided that the amino acid composition of the proteomes reflects the action of natural selection to enhance metabolic efficiency, synonymous codon usage bias as a measure of translation rates and shows increases in the abundance of less energetically costly amino acids in highly expressed proteins[31]. Comparison of the relative fixation rates of synonymous (silent) and nonsynonymous (amino acid-altering) mutations provides a means for understanding the mechanisms of molecular sequence evolution[36], and positive selection might be expected to follow viral emergence in a new host species because this event would obviously entail a major boost in the number of susceptible hosts and a concomitant increase in fitness[37]. The compiled codon usage data for different lineages of SARS-CoV-2 are presented in Supplementary Table 1. We summarize the synonymous codon usage of all different lineages of SARS-CoV-2 in Figure 2c and Supplementary Table 1. In the B.1.640.2 lineage, codon AGU was obviously biased for Ser, codon UUU was stronger for Phe in the B.1.640.1 and P.3 (Theta) lineages, and codon GCU was clearly biased for Ala in the B.1.617.2 (Delta) lineage. That is, a large selective difference between lineages of SARS-CoV-2 was required for strong codon usage bias.

### Clock-like prediction of focal outbreak points worldwide to provide warnings

To explain the epidemic outbreak points related to key mutations, we set the Epi-Clock device to predict potential epidemics and assist in the presentation of detailed mutation information for severely affected areas. Therefore, we analysed the whole evolutionary pathway in the timeline of lineages of SARS-CoV-2 presented in Supplementary Table 2; i.e., each lineage followed the same principle in which accumulations of mutations were the driving force of the phylogenetic relationships among different lineages of SARS-CoV, showing information for the earliest publicly collected samples in different regions of the world. At the same time, we summarized the smoothed distribution of new cases per million cases of SARS-CoV-2 in different severely affected areas, such as Africa, Asia, Europe, North America, Oceania, and South America, along the timeline from OWID[38] in Supplementary Figures 18-24. Interestingly, we found the pattern of an increasing number of other types of substitutions (shown with an asterisk) as the hitchhiking effect progressed, especially in the pre-breakout phase, even though some substitutions were replaced by other dominant genotypes, as shown in the red box in Figure 3a, Figure 3b, Supplementary Table 2 and Supplementary Figures 25-32.

**Fig. 3.**
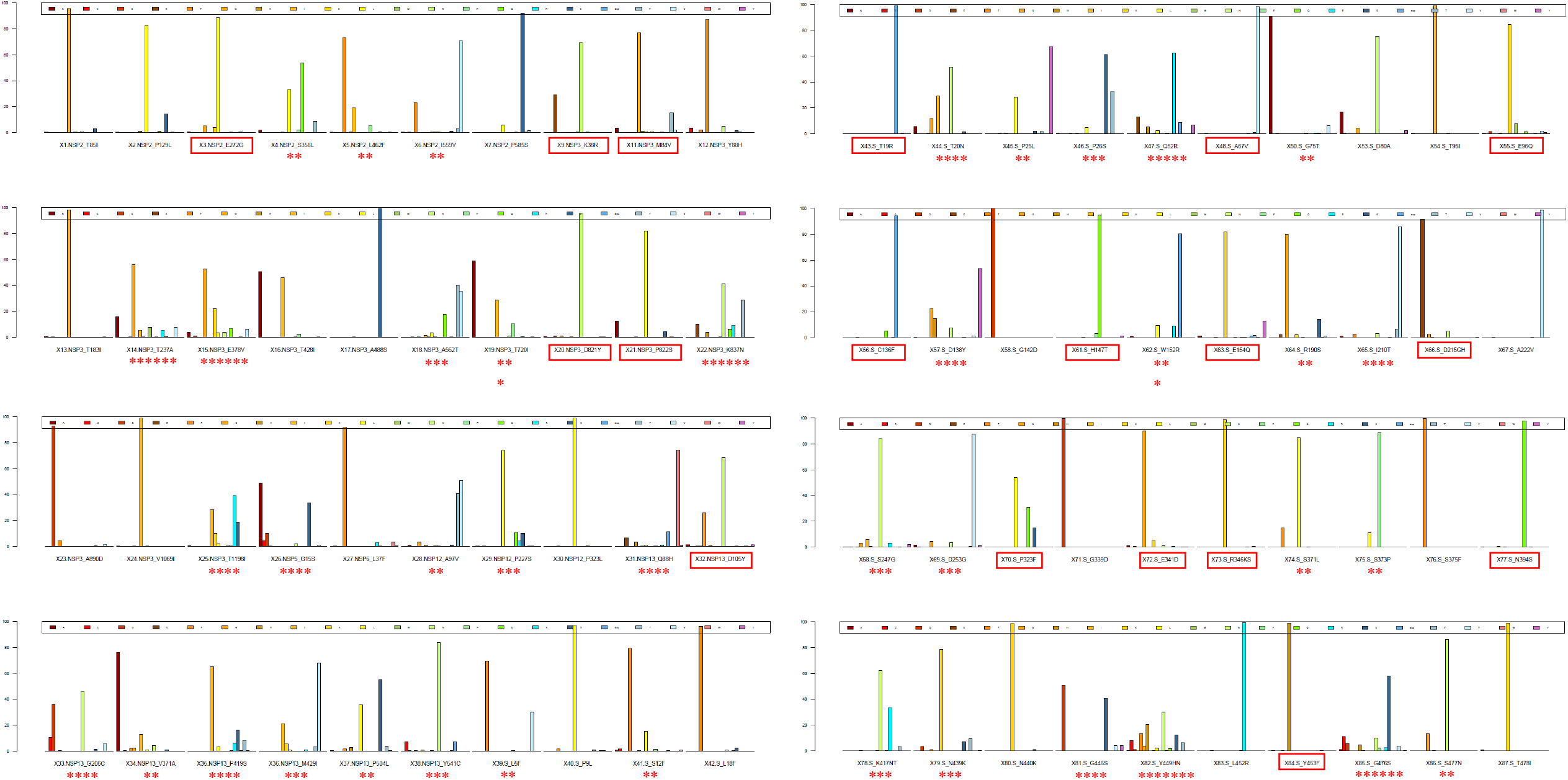
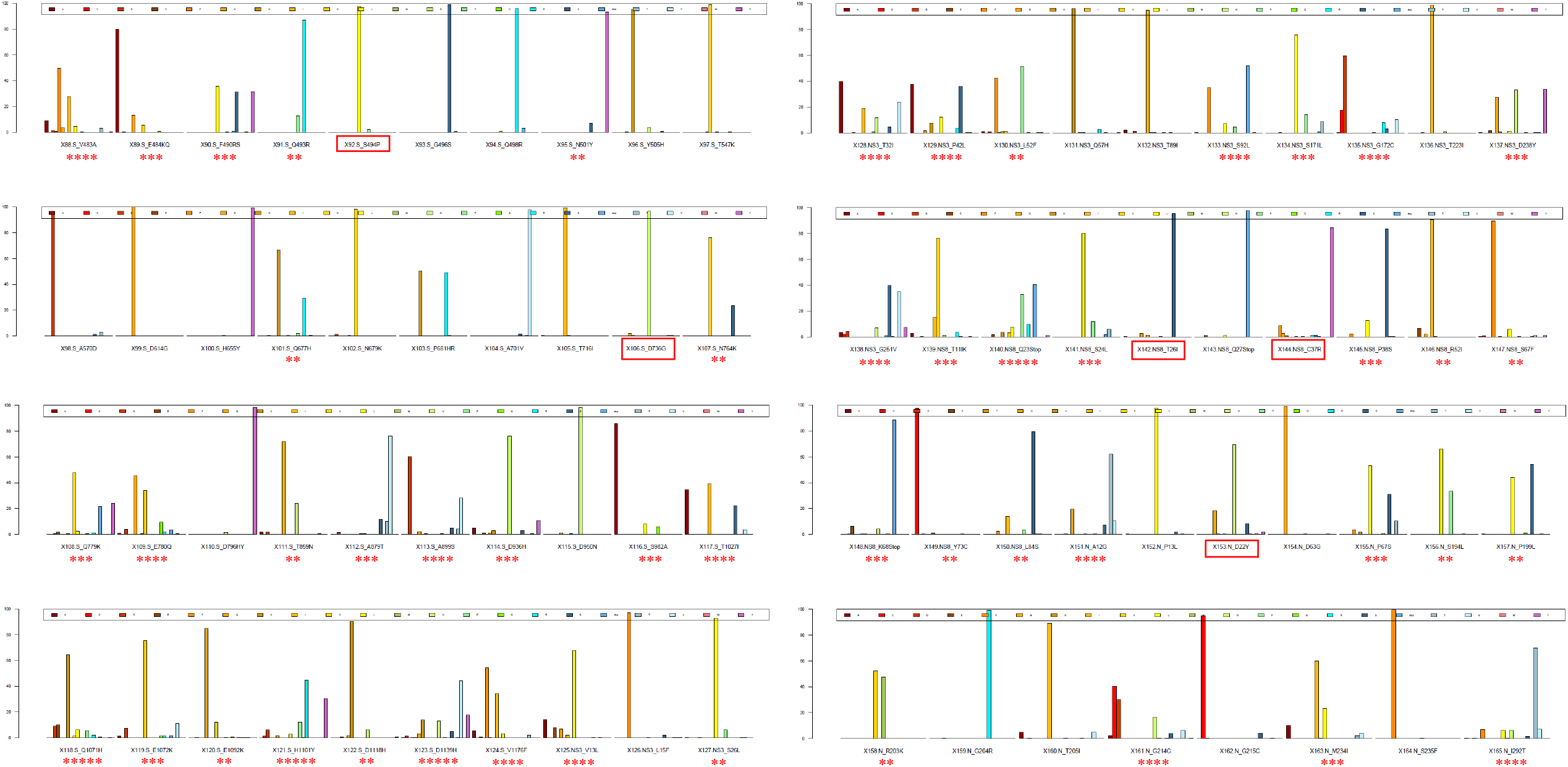
Illustration of mutation information for different lineages of SARS-CoV-2 in severely affected areas. **3a.**Frequency distribution of types of amino acid substitutions 1-87 in the pre-breakout phase. **3b**. Frequency distribution of types of amino acid substitutions 88-165 in the pre-breakout phase. Asterisks represent the pattern of an increasing number of other types of substitutions as the hitchhiking effect progressed, and red boxes show that some substitutions were replaced by other dominant genotypes recently.

We hypothesized that specific amino acid substitutions are triggers for outbreaks. To predict potential epidemic outbreaks, we proposed the ZHU algorithm presented in Figure 4a and tested it on different true sets of SARS-CoV-2 data before outbreaks to search for real significant mutational accumulation patterns related to the outbreak events. Phylogeographic inference methods may also be used to provide insights into the factors driving virus spread, and application of generalized linear modelling to explain Ebola virus migration rates between locations as a function of several potential predictors, to infer virus spread during West African Ebola outbreak, has been reported [26]. Geographic distances and population sizes at the location of origin and destination combined into a gravity model of spread, with virus transmission largely occurring within large population centres and geographic spread being more frequent over shorter distances[26]. However, our prediction of outbreak was only based on significant amino acids substitutes excluded other epi-factors. To avoid system errors, that is, errors caused by regions varying in other epi-factors such as population density, geographical environment, vaccination coverage rates and national/social rules and norms, we extracted the true sets by the control group in different individual countries or regions as the individual related values of the new cases per million. Then, we separately performed GLM analysis on individual countries/regions to find the optimal mutational patterns as in the training phase.

**Fig. 4.**
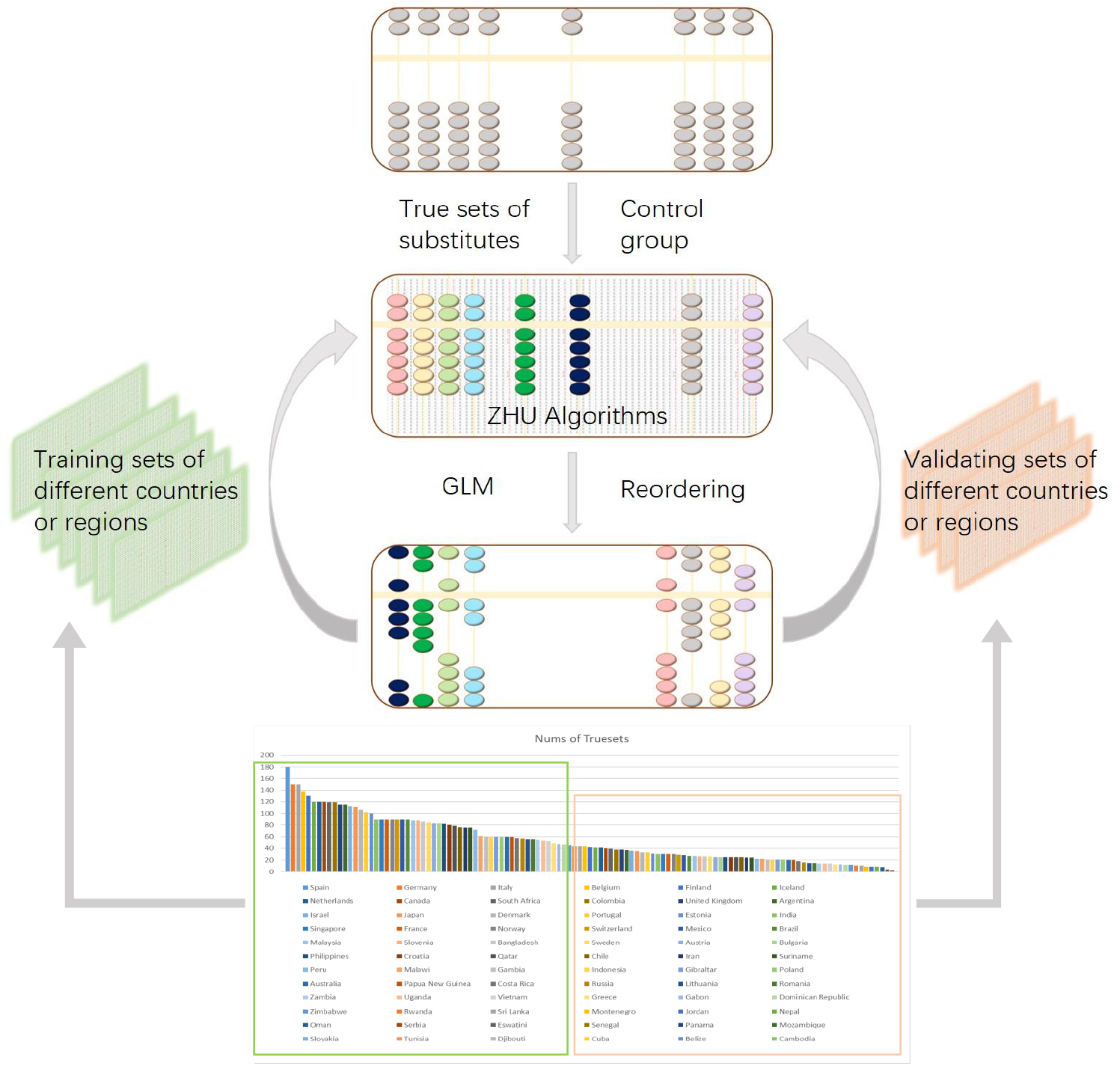
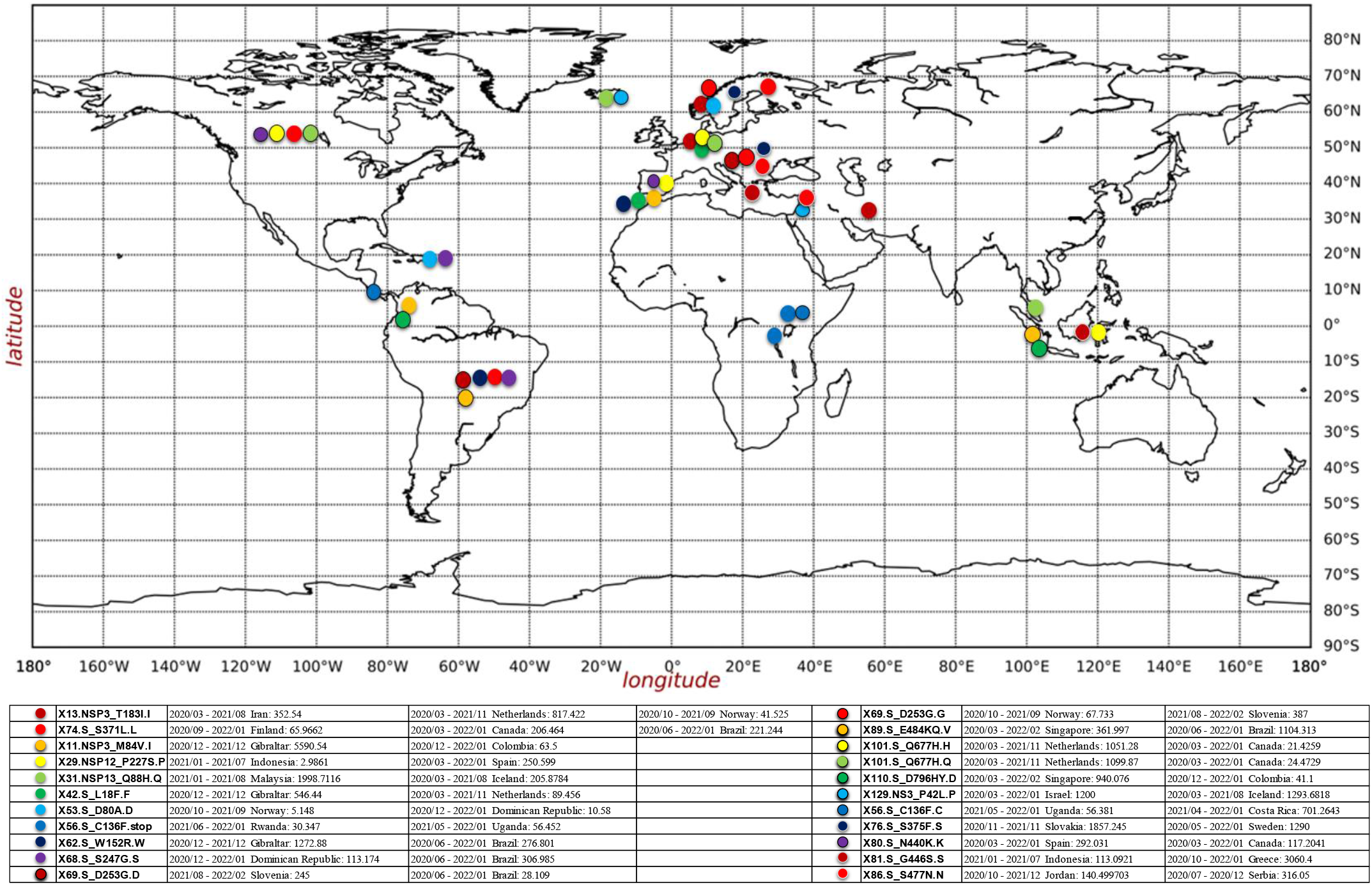
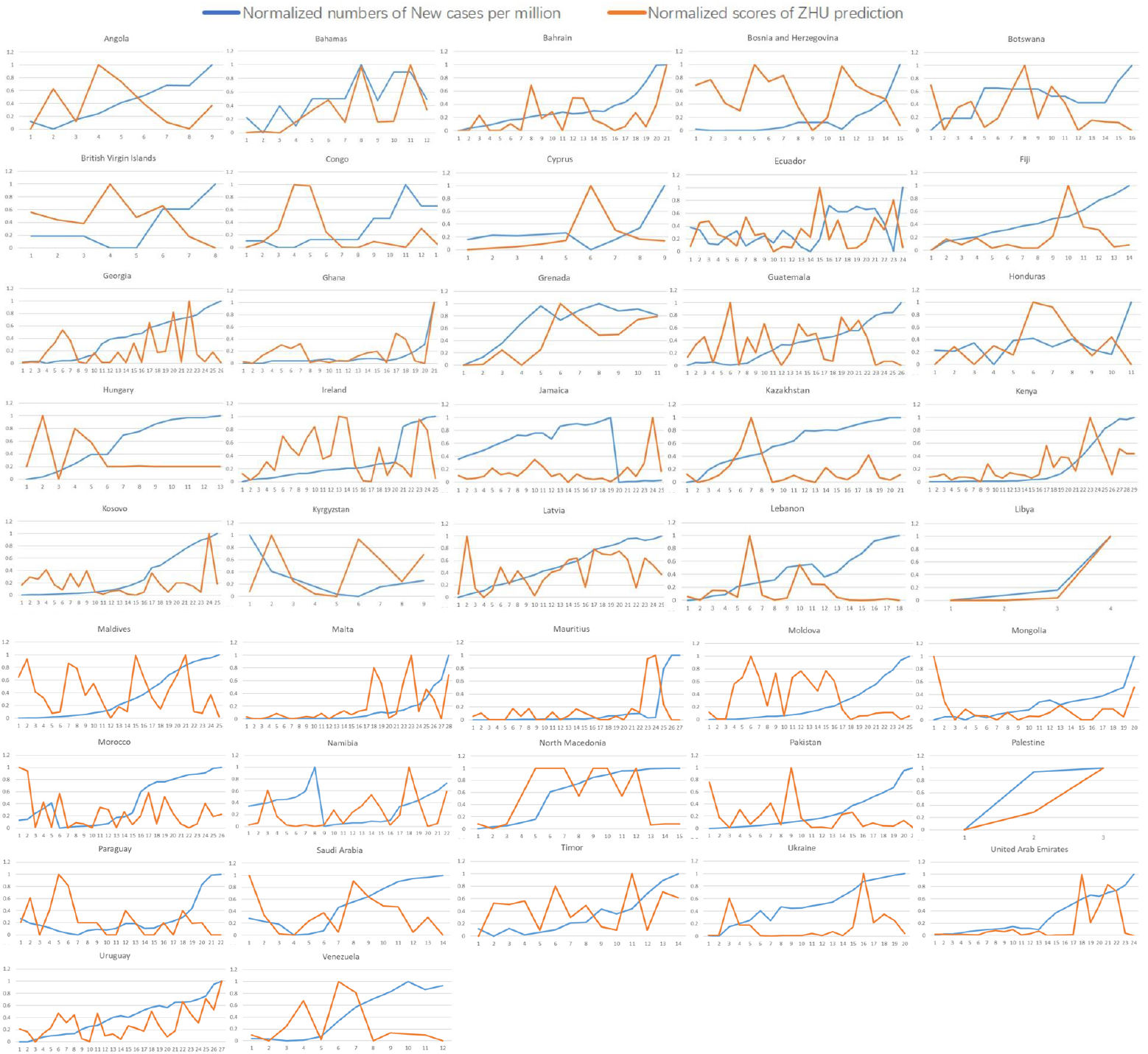
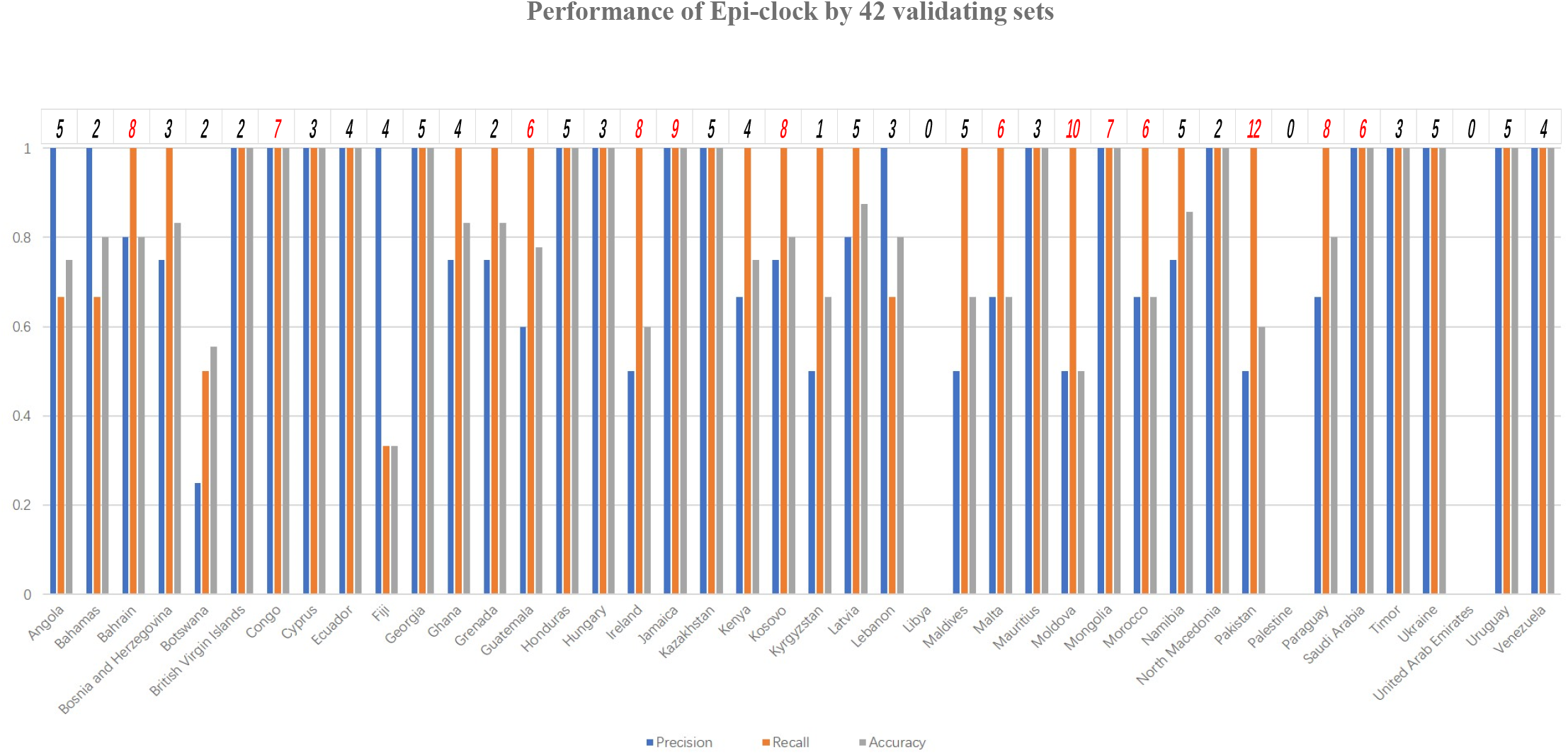
Algorithm and performance of Epi-Clock. **4a**. ZHU algorithm for inferring the timing of an epidemic of a new lineage related to increasing confirmed cases. **4b**. Prediction of 22 significant substitutions as the potential trigger for the outbreak from 75 training sets. **4c**. Performance of ZHU prediction measured by the relationship between observed confirmed numbers of new cases per million and normalized predicted numbers in different regions. **4d**. Performance of Epi-Clock evaluated by 42 validation sets.

Very interestingly, it was obvious that amino acid substitution type X13.NSP3_T183I.I was a potential epi-factor in Iran, the Netherlands and Norway. X74.S_S371L.L was significantly correlated with numbers of new cases in Finland, Canada and Brazil, as shown in Figure 4b. Across N generations of training by GLM and reordering, we found 171 significant substitutions as potential epi-factors within 55 different countries and regions, as demonstrated in Supplementary Table 3. Finally, we successfully presented the performance of the ZHU algorithm on 42 validation sets in Figure 4c. From most validations, we could accurately predict the potential pre-phase of the outbreak by ZHU prediction. Nevertheless, in only four countries, our model could not be fitted because of unavailable sequencing samples in the outbreak phase, such as in Jamaica, Kyrgyzstan, Libya, and Palestine. By counting the number of true instances, we summarized the precision, recall, and accuracy of Epi-Clock with the true validation sets as presented in Figure 4d and Supplementary Table 3, where the median interval before outbreaks was 5 days. As distinguished by EpiFactors [22], which included 815 proteins with 95 histones and protamines involved in epigenetic regulation, we extracted 171 significant amino acid substitutions as potential epi-factors within 55 different countries and regions, as presented in Supplementary Table 3.

## Discussion

The epidemic viruses did not spread into neighbouring countries showing susceptibility to significant outbreaks but a lower risk of introduction, which revealed the large epidemic to be a heterogeneous and spatially dissociated collection of transmission clusters of varying size, duration and connectivity[39]. Through the analysis of over 900 full genomes from an epidemiological collection of more than 26000 strains from Australia and New Zealand, fundamental differences in the phylodynamics of two co-circulating lineages of influenza B virus were determined by examining the complex relationships between virus transmission, age of infection and receptor binding preference, which are new factors that are important determinants of influenza B evolution and epidemiology[40]. In short, parallel adaptation occurs in the context of several viral behaviours, including cross-species transmission, drug resistance, and host immune escape, and its existence suggests that at least some aspects of virus evolution and emergence are repeatable and predictable[41]. The development of a combination of pathogenic epidemic prediction and pathogenic testing strategies not only improved the containment of COVID-19 but also contributed to the control of other infectious diseases, such as HIV and human papillomavirus, and non-communicable diseases[42].

Rarely, viruses gain the ability to spread efficiently within a new host that was not previously exposed or susceptible, which involves either increased exposure or the acquisition of variations that allow them to overcome barriers to infection of the new host. It is well known that host switching leads to viral emergence, considering the evolutionary mechanisms, virus‒host interactions, host range barriers to infection, and processes that allow efficient host-to-host transmission in the new host population[14]. For instance, at least 49 human-to-swine transmission events occurred globally during 2009-2011, highlighting the ability of the H1N1pdm09 virus to transmit repeatedly from humans to swine, even following adaptive evolution in humans[43]. Whether hosts and their symbionts speciate in parallel, by co-speciation, or through host shifts is a key issue in host-symbiont evolution. Convincing cases of co-speciation are rare (7%), and examining the relationships between short-term co-evolutionary dynamics and long-term patterns of diversification in host-symbiont associations revealed that these events could occur following host shifts and do not necessarily involve co-speciation. Therefore, there is now substantial evidence that the co-evolutionary dynamics of hosts and parasites do not favour long-term co-speciation[44]. Above all, host range is a viral property reflecting natural hosts that are infected either as part of a principal transmission cycle or, less commonly, via spillover infections into alternative hosts. Wildlife host species richness has been proposed as an important predictor of disease emergence[11].

Host immunity is a major driver of pathogen evolution and a major determinant of pathogen diversity; however, the complexity of many host‒pathogen interactions is dynamically important, broadening the definition of a pathogen’s immunological phenotype or what can be thought of as its immunological niche[45]. Recently, it was reported that a higher rate of severe outcomes and considerable mortality exist in unvaccinated people, especially older adults. That is, the strict and comprehensive pandemic control strategies implemented in Shanghai were able to reduce the number of people infected and to support early diagnosis and appropriate treatment for severe COVID-19 so that the case fatality rate could be minimized and to buy time for full vaccination coverage[46]. Of course, our study has limitations. The advent of high-content single-cell technologies has facilitated a greater understanding of the properties of host cells harbouring infection, the host’s pathogen-specific immune responses, and the mechanisms pathogens have evolved to escape host control[47] [48]. It is important to identify natural genetic variants that underlie variation in the host innate immune response to infection and analyse the mechanisms by which such variants alter these responses[49]. It was considered that mutation, natural selection, and genetic drift combined to shape codon usage patterns with different strength proportions. Natural selection causes micro-evolutionary changes that maintain or increase the fitness competence for the organism, while random gene drift considers the frequency of a gene is fixed by the population and is strongly linked to their size without taking into account their effect in the organism phenotype[50]. Random genetic drift plays a role in the processes of transcription, translation, replication[32]. Unlike other RNA viruses, such influenza, SARS-CoV-2 has a genetic proofreading mechanism regulated by NSP14 and NSP12 (a.k.a RNA-dependent RNA polymerase)[51, 52], that enables to have a higher fidelity in its replication. However, the host gene editing has been found to be the major source for existing SARS-CoV-2 mutation counting for 65% of reported mutations[53]. Meanwhile, it provided important insights into how natural selection has shaped immunity and host defence genes in specific human populations and in the human species as a whole, information that is helping delineate genes that were important for host defence and increase our understanding of how past selection had an impact on disease susceptibility in modern populations[34].

## Conclusions

Above all, Omicron strains are the turning point of SARS-CoV-2 epidemic with strong infectivity, which provide great opportunity to the viral adaptive evolution within large scales of individual population size. It represents diverse amino acid substitutes on different sites achieved by different ages, species, or physiological environment. Except with mutational shifts among different hosts, pathogenicity and fatality have come down gradually. We believe that it would never repeat serious clinical events again only according to nature selection. Because SARS-CoV-2 has been totally adaptive in human body. With the accumulation of sequencing datasets, the performance of our strategy will improve, and the approach will become more sensitive in its ability to predict the potential trigger of epidemic outbreaks, which will ultimately facilitate responses to future outbreaks of concern.

## Data Availability

All data produced in the present study are available upon reasonable request to the authors

https://bioinfo.liferiver.com.cn

## Abbreviations

DNA: Deoxyribonucleic acid
RNA: Ribonucleic acid
vRNP: viral ribonucleoprotein
SARS-CoV-2: Severe acute respiratory syndrome coronavirus 2
RBD: Receptor-binding domain
EBOV: Ebola virus
NCLDV: Nucleocytoplasmic large DNA virus
MVs: *Marseilleviridae*
DG: D614G substitute
MEGA11: Molecular Evolutionary Genetics Analysis
RSCU: Relative synonymous-codon usage
OWID: Our World in Data
GLM: Generalized linear model
TP: True positive
FP: False positive
FN: False negative
TN: True negative
SARS-CoV: Severe acute respiratory syndrome-related coronavirus
SADS: Swine acute diarrhoea syndrome coronavirus
NL63: Human coronavirus NL63
MERS: Middle East respiratory syndrome-related coronavirus
London1: Betacoronavirus England 1
HKU5: Bat coronavirus HKU5
HKU4: Bat coronavirus HKU4
HKU3: Bat coronavirus HKU3
HKU2: Bat coronavirus HKU2
BATS: Bat SARS-like coronavirus WIV1
OC43: Human coronavirus OC43
HKU9: Rousettus bat coronavirus HKU9
HKU1: Human coronavirus HKU1
229E: Human coronavirus 229E
ORF: Open reading frame
NSP: Non-structural protein
S: Surface glycoprotein
E: Envelope protein
M: Membrane glycoprotein
N: Nucleocapsid phosphoprotein
HIV: Human immunodeficiency virus
H1N1pdm09: 2009 H1N1 Pandemic

## Declarations

### Ethical approval and consent to participate

All protocols were approved by the Liferiver Science and Technology Institute, Shanghai ZJ Bio-Tech Co., Ltd and Use Committee (Shanghai, China).

### Consent for publication

Not applicable.

### Availability of data and material

All shared mutations and substitutes from different hosts, lineages or regions are available in the supplementary materials and https://bioinfo.liferiver.com.cn/#/home.

### Author contributions

The authors’ responsibilities were as follows: JBS and CJ designed and conducted the research; CJ analysed the data and performed the analysis; CJ wrote the paper; JBS and CJ revised the manuscript; CJ had primary responsibility for the final content. All authors read and approved the final manuscript.

### Corresponding author

Correspondence to Cong Ji and Junbin (Jack) Shao.

## Acknowledgements

Appreciation goes to Chen Han and Wang Guangzhong for their valuable suggestions and comments on this work. Thank you so much for the discussion from Chen Si, Liu Yan, and Zhang Hanyan. Many facets of the user-interface design benefited from help by Niu Xingsheng, Pan Yajie and Lu Wang. Great thanks to Elizabeth Sung for flow and language corrections. All the other data supporting the findings of this study and the computational code used in this study are available from the corresponding authors upon reasonable request. The other authors declare no competing interests.

