## Supplementary Figure1-32 for "Epi-Clock: A sensitive platform to help understand pathogenic disease outbreaks and facilitate the response to future outbreaks of concern"

**Supplementary Figure 1. Intra-species and inter-species genetic distances of *Coronaviridae*.** *Coronaviridae* include SARS-CoV-2, SARS-CoV, SADS, OC43, NL63, MERS, London1, HKU9, HKU5, HKU4, HKU3, HKU2, HKU1, BATS, and 229E.

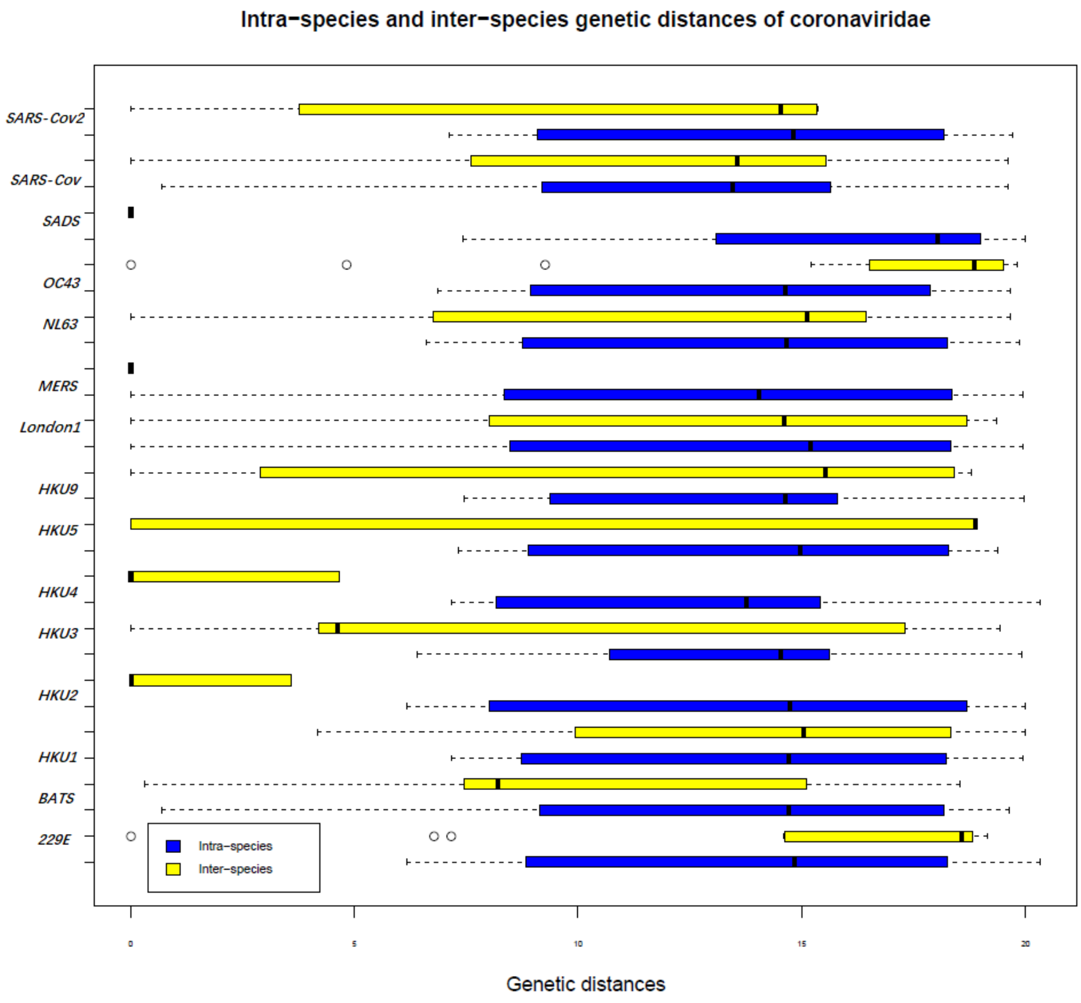

**Supplementary Figure 2. Atlas of new ages within different populations.** Here, we separately present the divergence of species or subspecies from SARS-CoV-2, i.e., SARS-CoV, SADS, OC43, NL63, MERS, London1, HKU9, HKU5, HKU4, HKU3, HKU2, HKU1, BATS and 229E. Red represents insertions, and blue represents deletions.

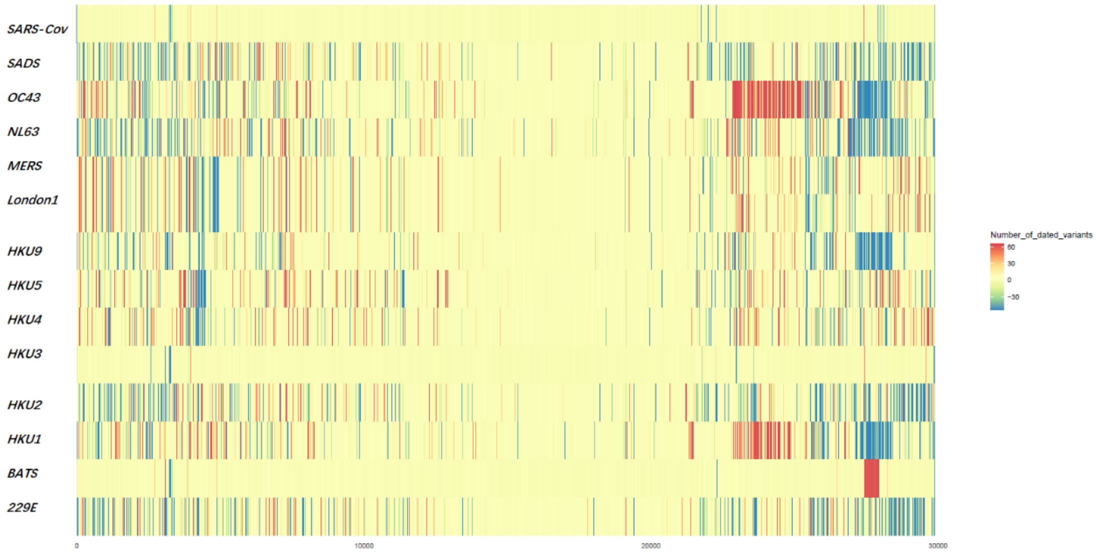

**Supplementary Figure 3. The indels of whole genome sequences of human coronavirus 229E between different hosts.** Several hosts are shown, such as (from outside to inside) *Camelus*, *Hipposideros*, *Macronycteris vittata*, and *Vicugna pacos*, while the genome of *Homo sapiens* was used as the reference genome.

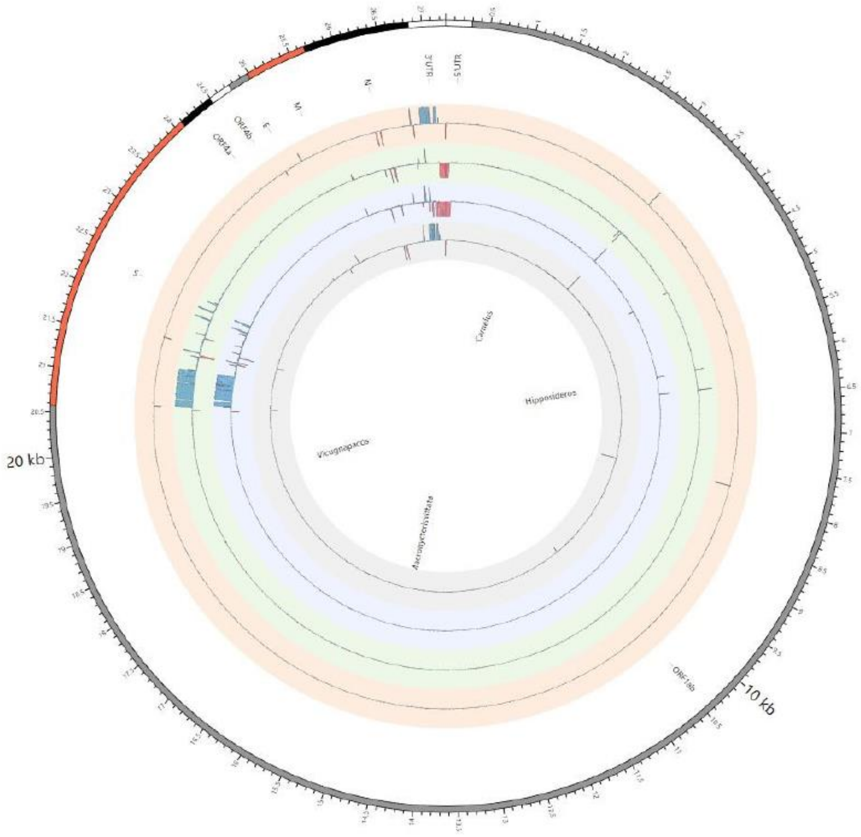

**Supplementary Figure 4. The indels of whole genome sequences of Rhinacovirus between different hosts.** Several hosts are shown, such as (from outside to inside) *Rhinolophus ferrumequinum* and *Rhinolophus affinis*, while the genome of *Sus scrofa* was used as the reference genome.

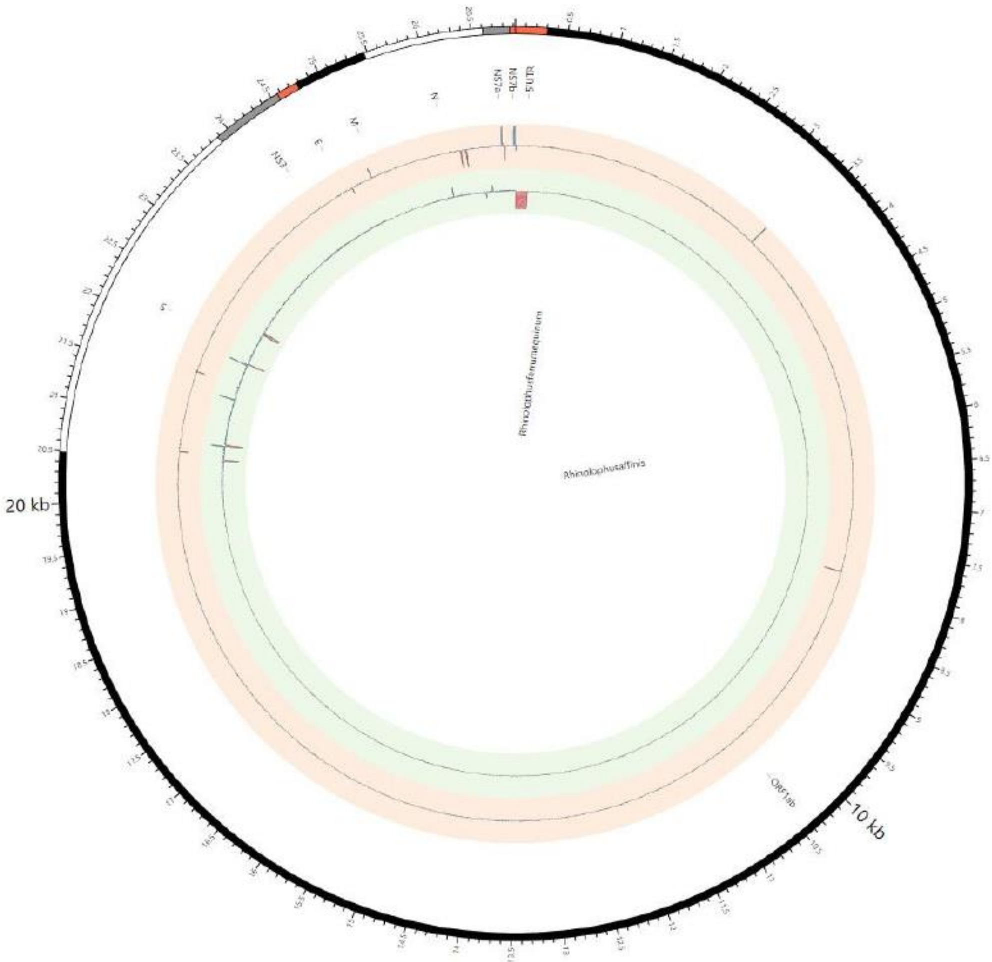

**Supplementary Figure 5. The indels of whole genome sequences of Tegacovirus between different hosts.** Several hosts are shown, such as (from outside to inside) *Felis catus*, *Canis lupus familiaris*, and Feliformia, while the genome of *Sus scrofa* was used as the reference genome.

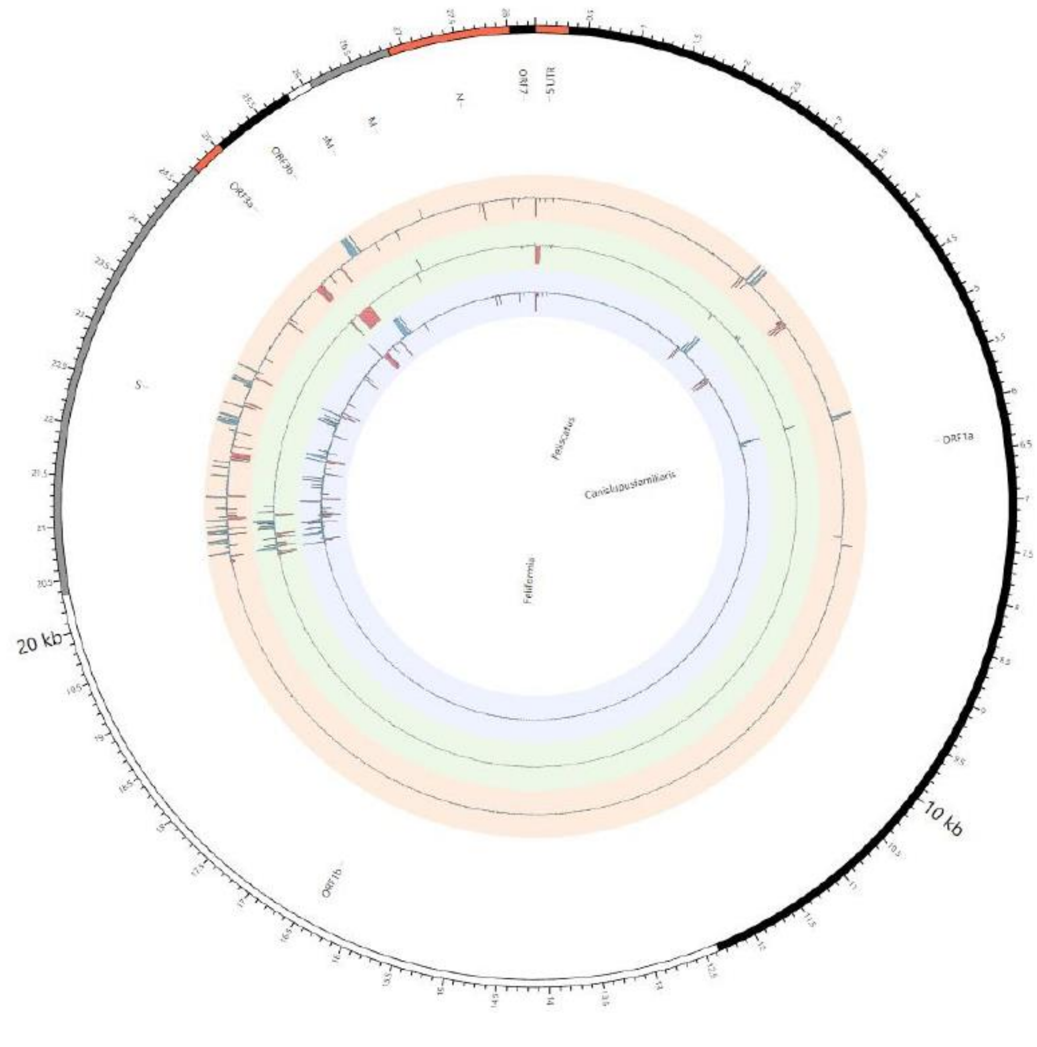

**Supplementary Figure 6. The indels of whole genome sequences of human coronavirus OC43 between different hosts.** Several hosts are shown, such as (from outside to inside) *Bos taurus*, *Bos grunniens*, Canis lupus familiaris, *Pan troglodytes verus*, *Sus scrofa*, *Hydropotes inermis*, *Bubalus bubalis*, *Camelus bactrianus*, *Equus caballus*, *Kobus ellipsiprymnus*, *Odocoileus virginianus*, *Rusa unicolor*, Bovidae, *Giraffa camelopardalis*, and *Vicugna pacos*, while the genome of *Homo sapiens* was used as the reference genome.

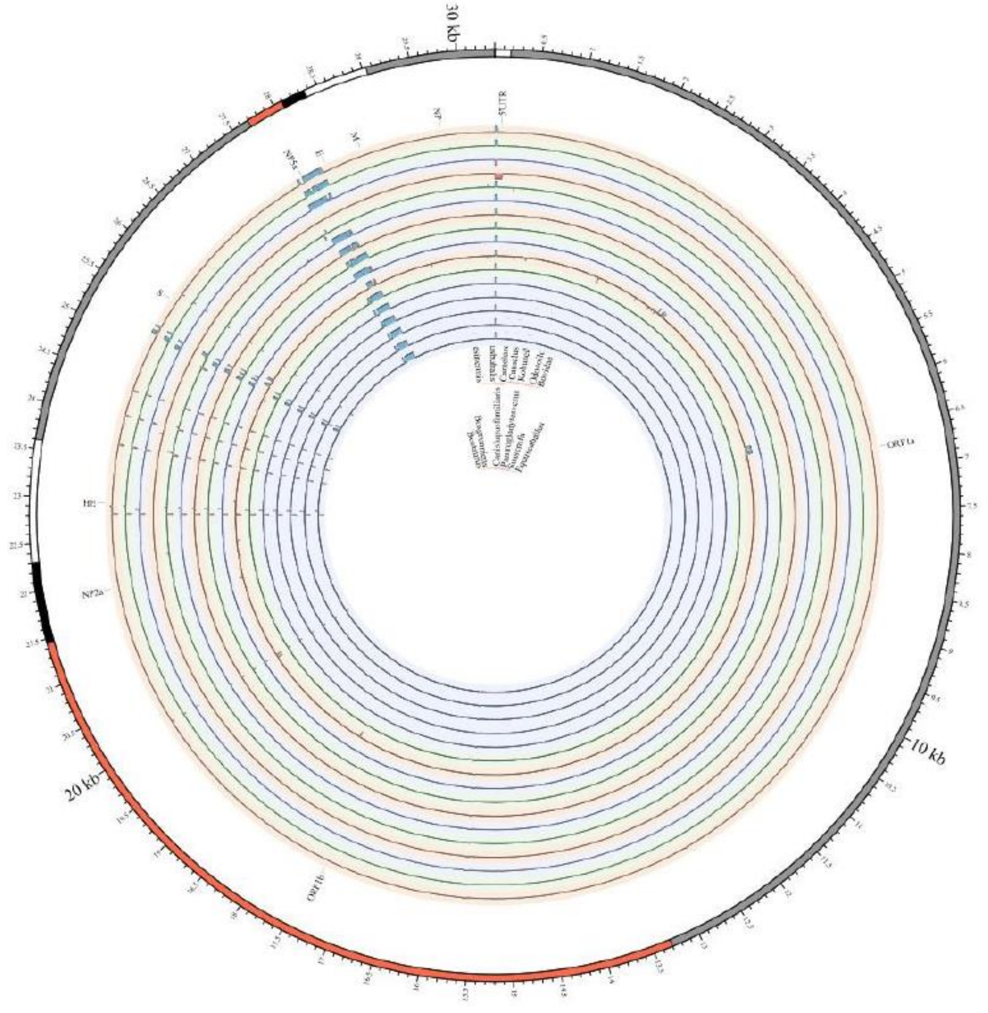

**Supplementary Figure 7. The indels of whole genome sequences of Middle East respiratory syndrome-related coronavirus between different hosts.** Several hosts are shown, such as (from outside to inside) *Camelus dromedaries*, *Lama glama*, *Neoromicia capensis*, *Hypsugo savii*, *Pipistrellus kuhlii*, and *Vespertilio sinensis*, while the genome of *Homo sapiens* was used as the reference genome.

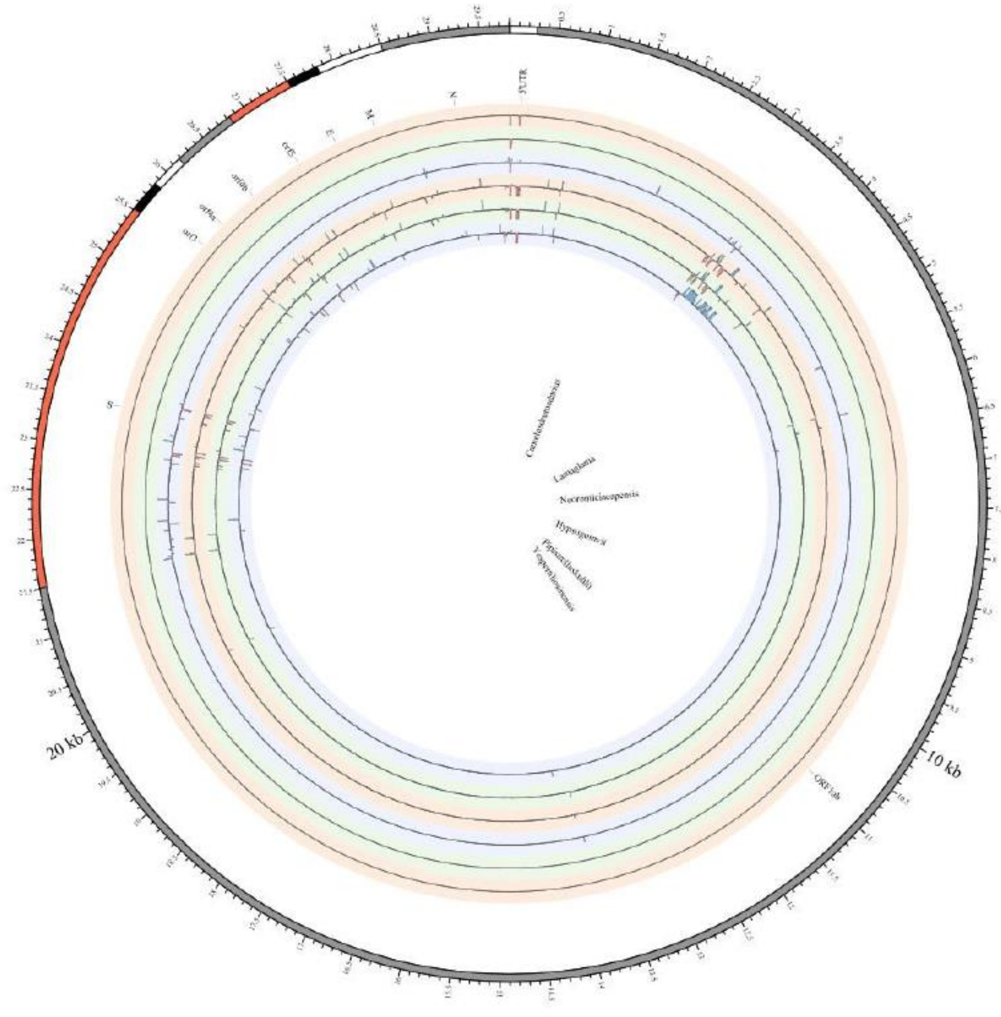

**Supplementary Figure 8. The indels of whole genome sequences of severe acute respiratory syndrome coronavirus 2 between different hosts.** Several hosts are shown, such as (from outside to inside) *Mus musculus*, *Rhinolophus sinicus*, *Mustela lutreola*, *Rhinolophus macrotis*, *Canis lupus familiaris*, *Panthera tigris jacksoni*, *Rhinolophus affinis*, *Aselliscus stoliczkanus*, *Rhinolophus ferrumequinum*, Chiroptera, *Rhinolophus pusillus*, *Chaerephon plicatus*, *Chlorocebus aethiops*, *Paradoxurus hermaphroditus*, Viverridae, *Paradoxurus hermaphroditus*, and *Paguma larvata*, while the genome of *Homo sapiens* was used as the reference genome.

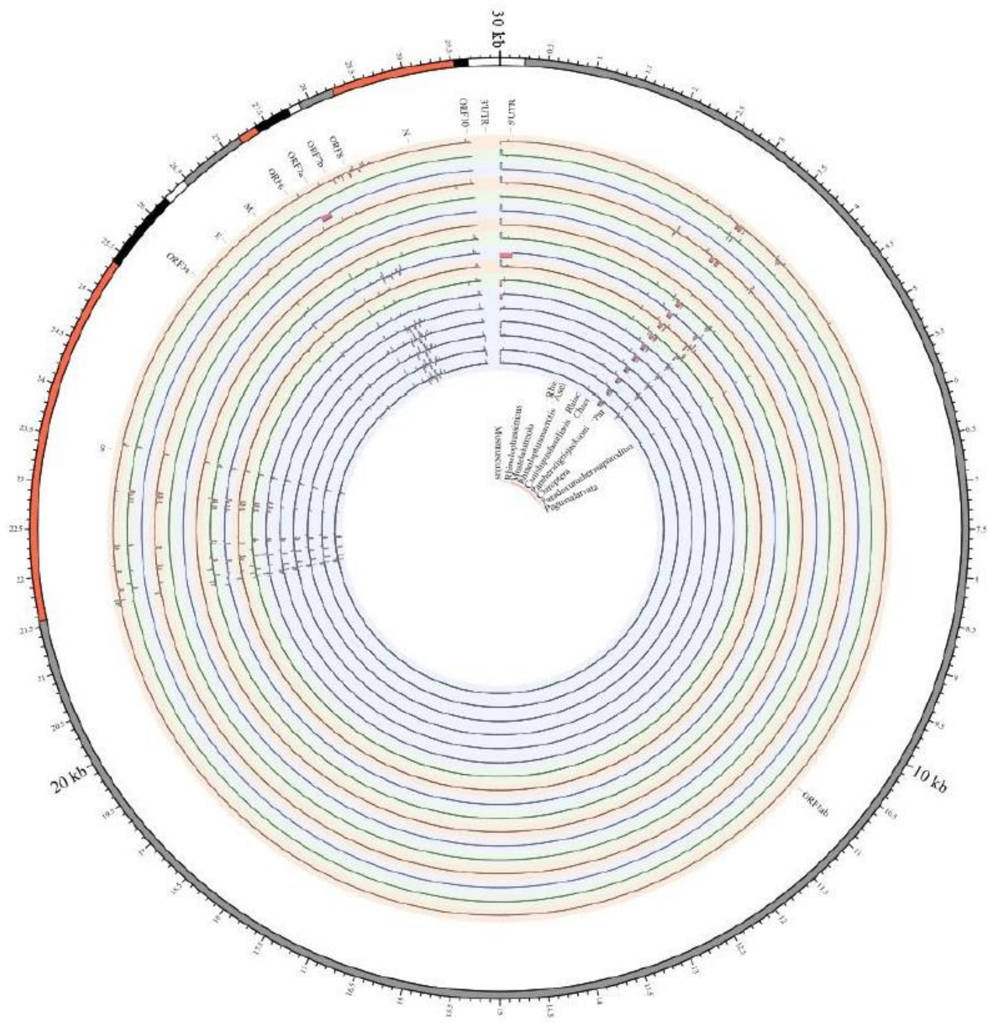

**Supplementary Figure 9. The indels of whole genome sequences of Deltacoronavirus between different hosts.** Several hosts are shown, such as (from outside to inside) *Pycnonotus jocosus*, *Mareca*, *Lonchura striata*, Zosteropidae, *Falco*, *Chlamydotis*, Columbidae, Galliformes, Passeridae, Muscicapidae, *Turdus hortulorum*, Ardeidae, *Gallinula chloropus*, and *Coturnix japonica*, while the genome of *Sus scrofa* was used as the reference genome.

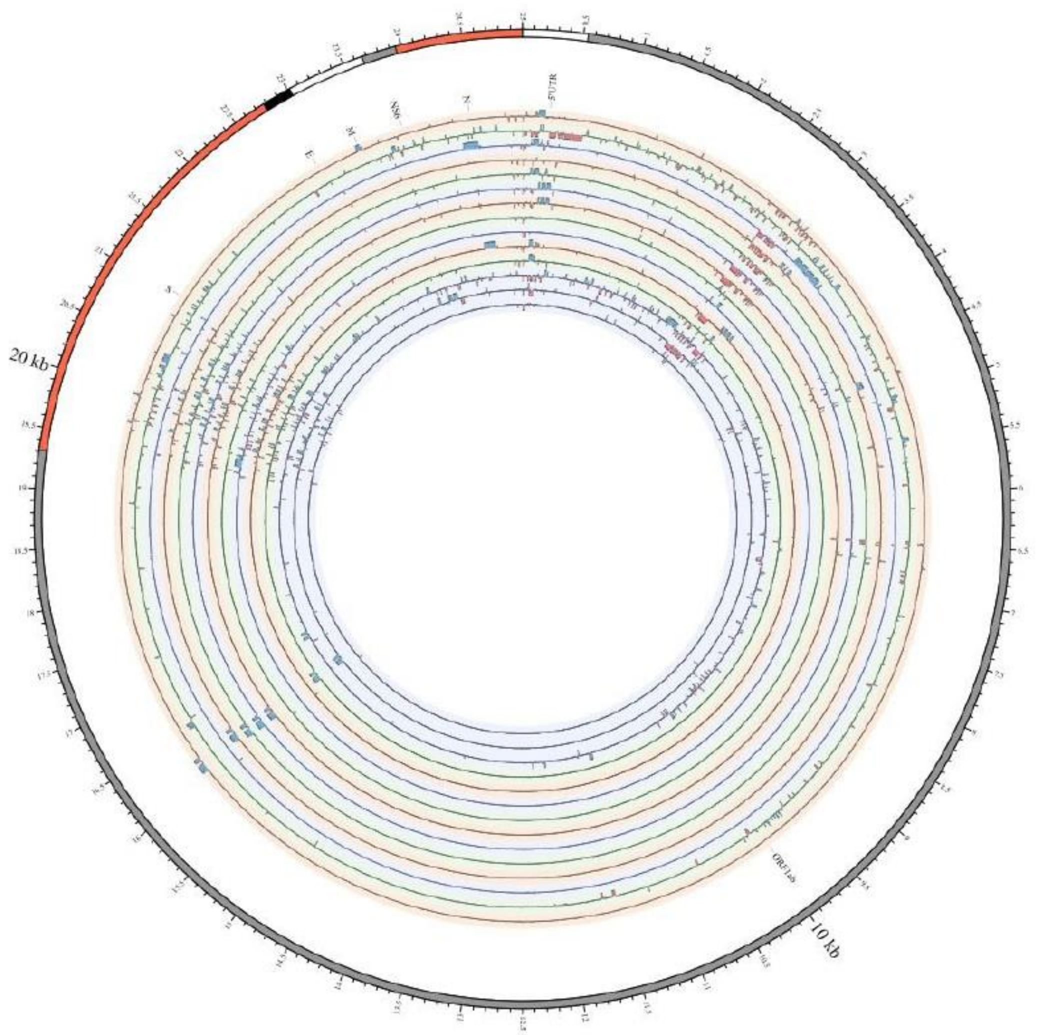

**Supplementary Figure 10. The indels of whole genome sequences of avian coronavirus between different hosts.** Several hosts are shown, such as (from outside to inside) Phasianinae, Anatidae, and *Meleagris gallopavo*, while the genome of *Gallus gallus* was used as the reference genome.

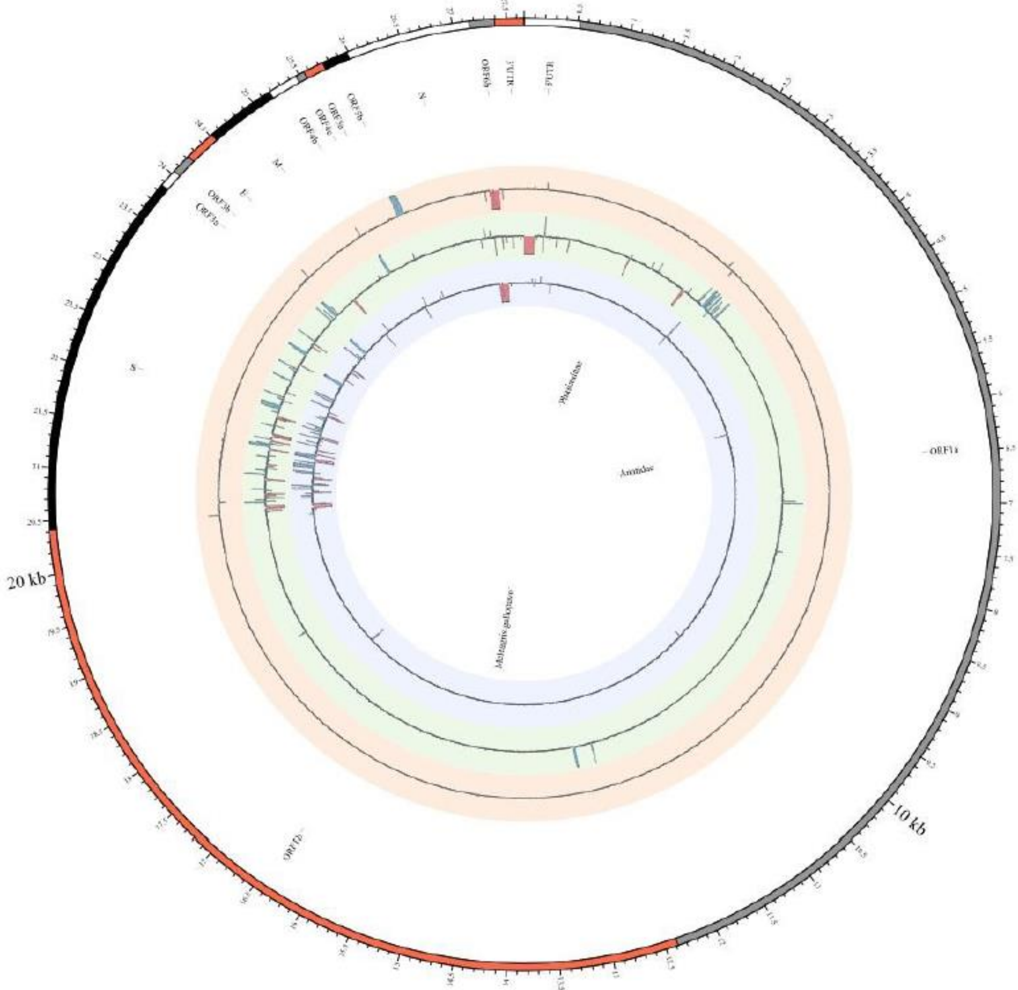

**Supplementary Figure 11. The whole-genome percentage nucleotide identity of different lineages of SARS-CoV-2.** There were several lineages such as P.3(Theta), P.1(Gamma), C.37(Lambda), C.36.3, C.1.2, B.1.640.2, B.1.640.1, B.1.621(Mu), B.1.617.3, B.1.617.2(Delta), B.1.617.1(Kappa), B.1.526(Lota), B.1.525(Eta), B.1.351(Beta), B.1.1.7(Alpha), B.1.1.529(Omicron), and B.1.1.318.

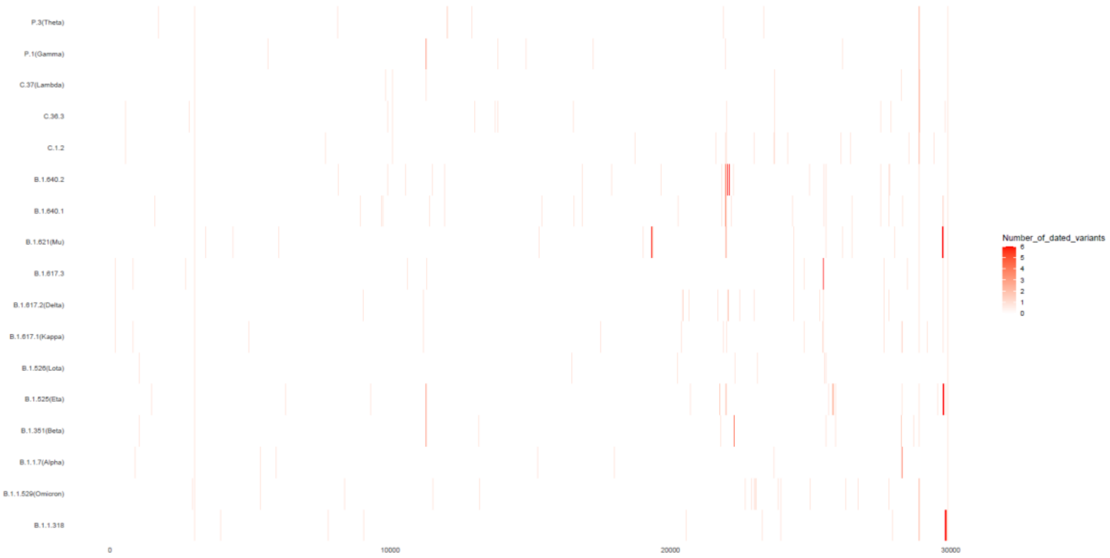

**Supplementary Figure 12. The richness distribution of the mutation rate of C->T across the whole SARS-CoV-2 genome in different lineages.** There were several lineages such as P.3(Theta), P.1(Gamma), C.37(Lambda), C.36.3, C.1.2, B.1.640.2, B.1.640.1, B.1.621(Mu), B.1.617.3, B.1.617.2(Delta), B.1.617.1(Kappa), B.1.526(Lota), B.1.525(Eta), B.1.351(Beta), B.1.1.7(Alpha), B.1.1.529(Omicron), and B.1.1.318.

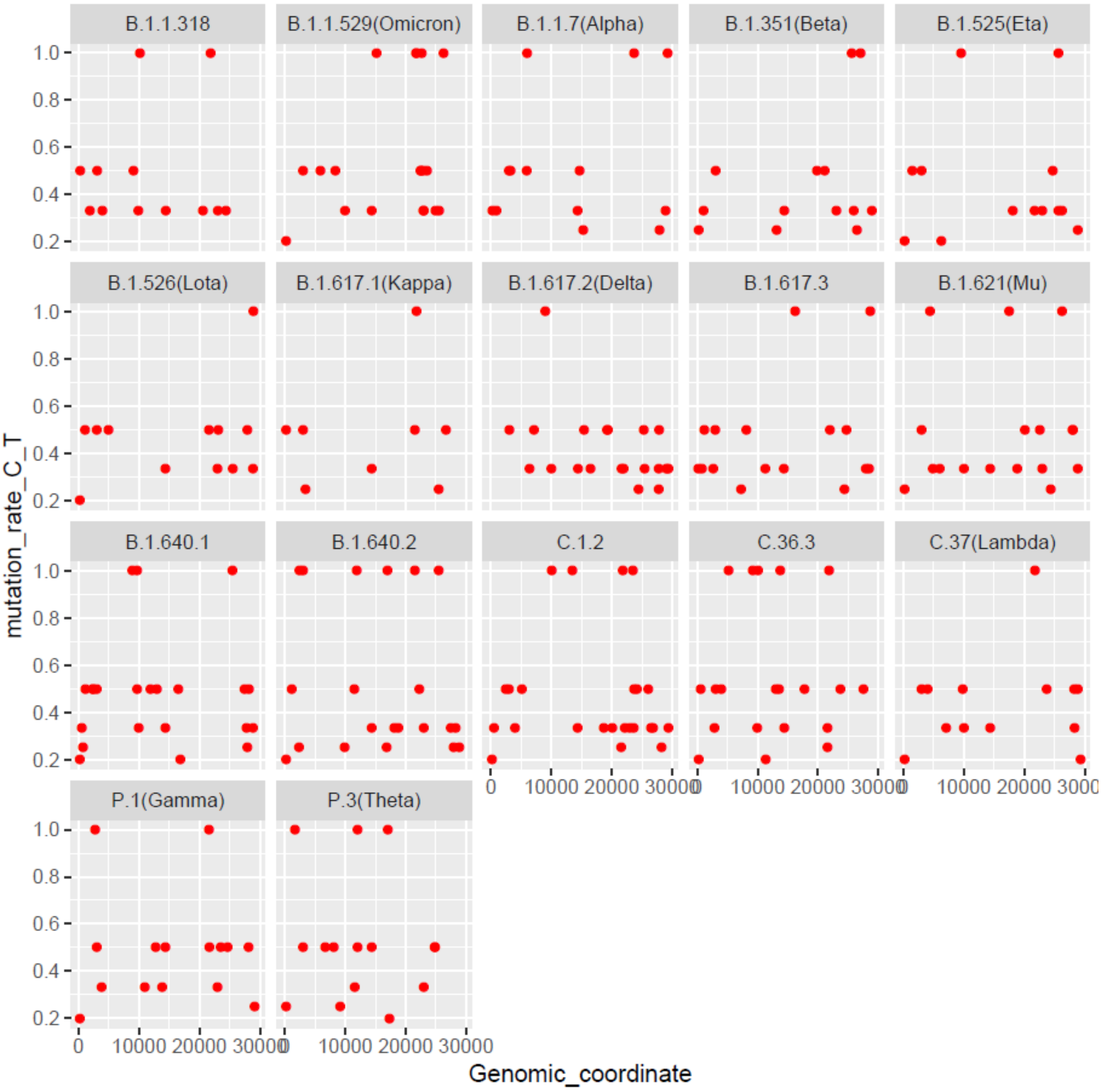

**Supplementary Figure 13. The richness distribution of the mutation rate of G->T across the whole SARS-CoV-2 genome in different lineages.** The same as above Supplementary Figure 12.

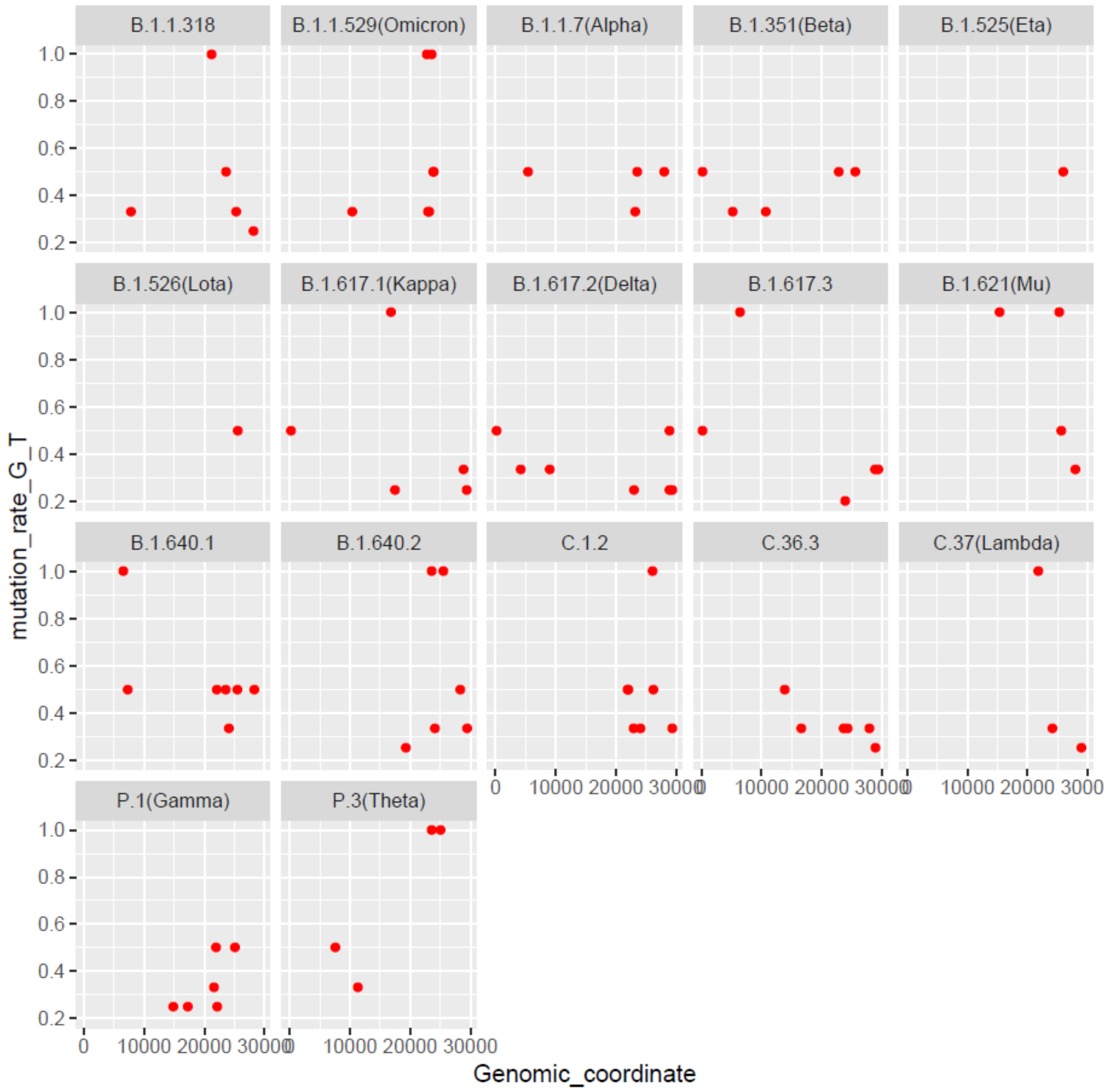

**Supplementary Figure 14. The richness distribution of the mutation rate of A->T across the whole SARS-CoV-2 genome in different lineages.** The same as above Supplementary Figure 12.

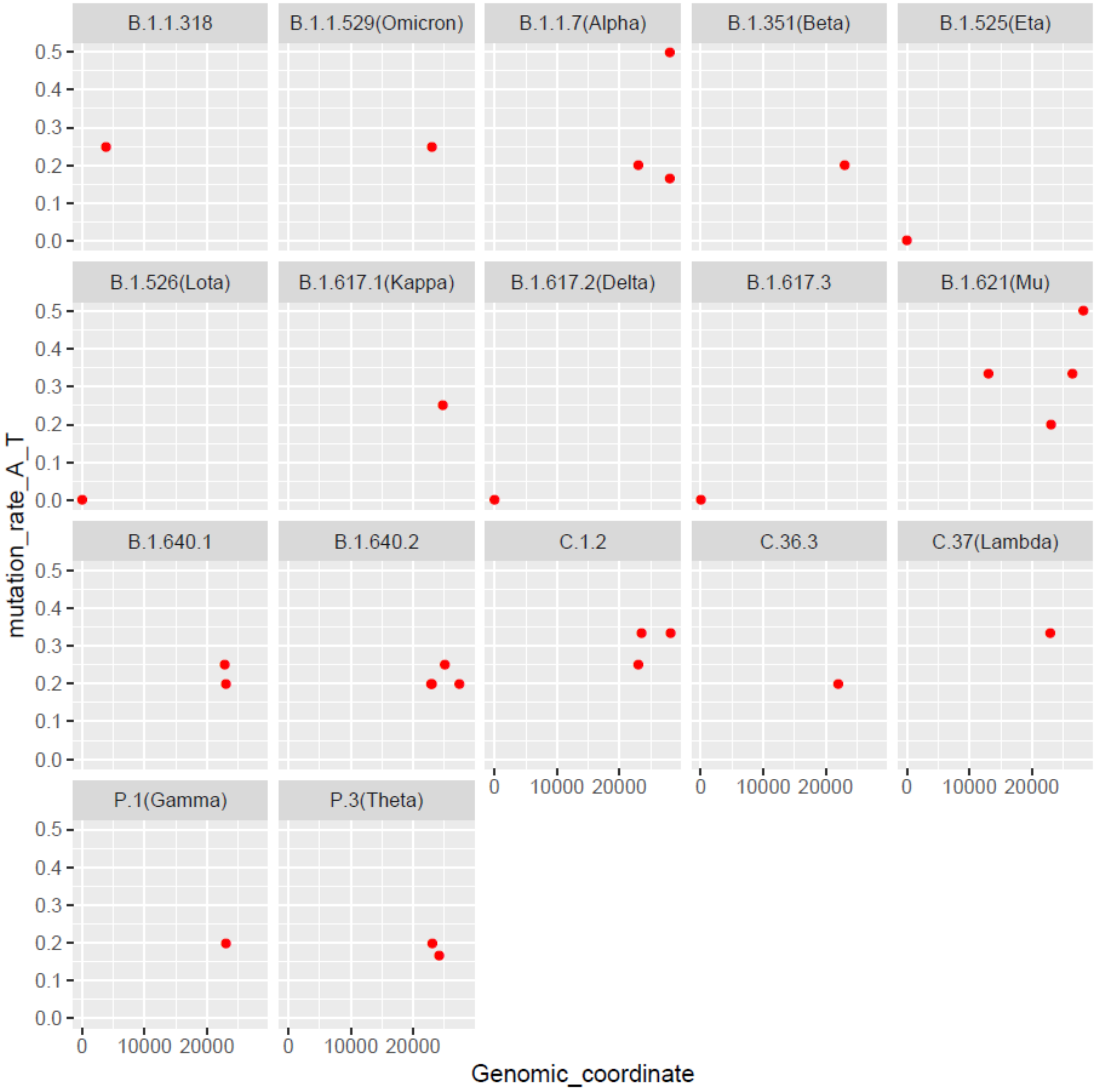

**Supplementary Figure 15. The richness distribution of the mutation rate of T->C across the whole SARS-CoV-2 genome in different lineages.** The same as above Supplementary Figure 12.

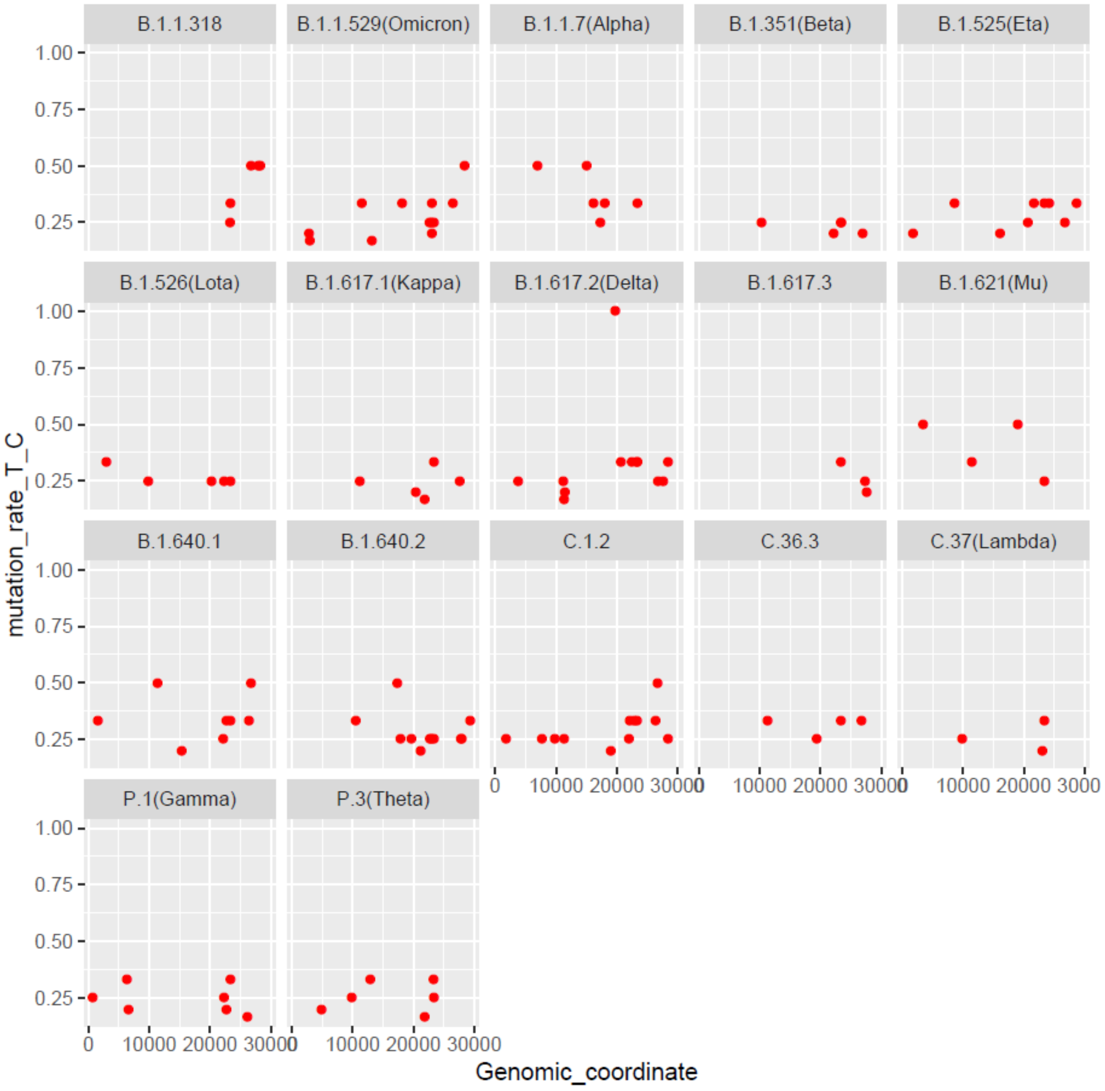

**Supplementary Figure 16. The richness distribution of the mutation rate of T->G across the whole SARS-CoV-2 genome in different lineages.** The same as above Supplementary Figure 12.

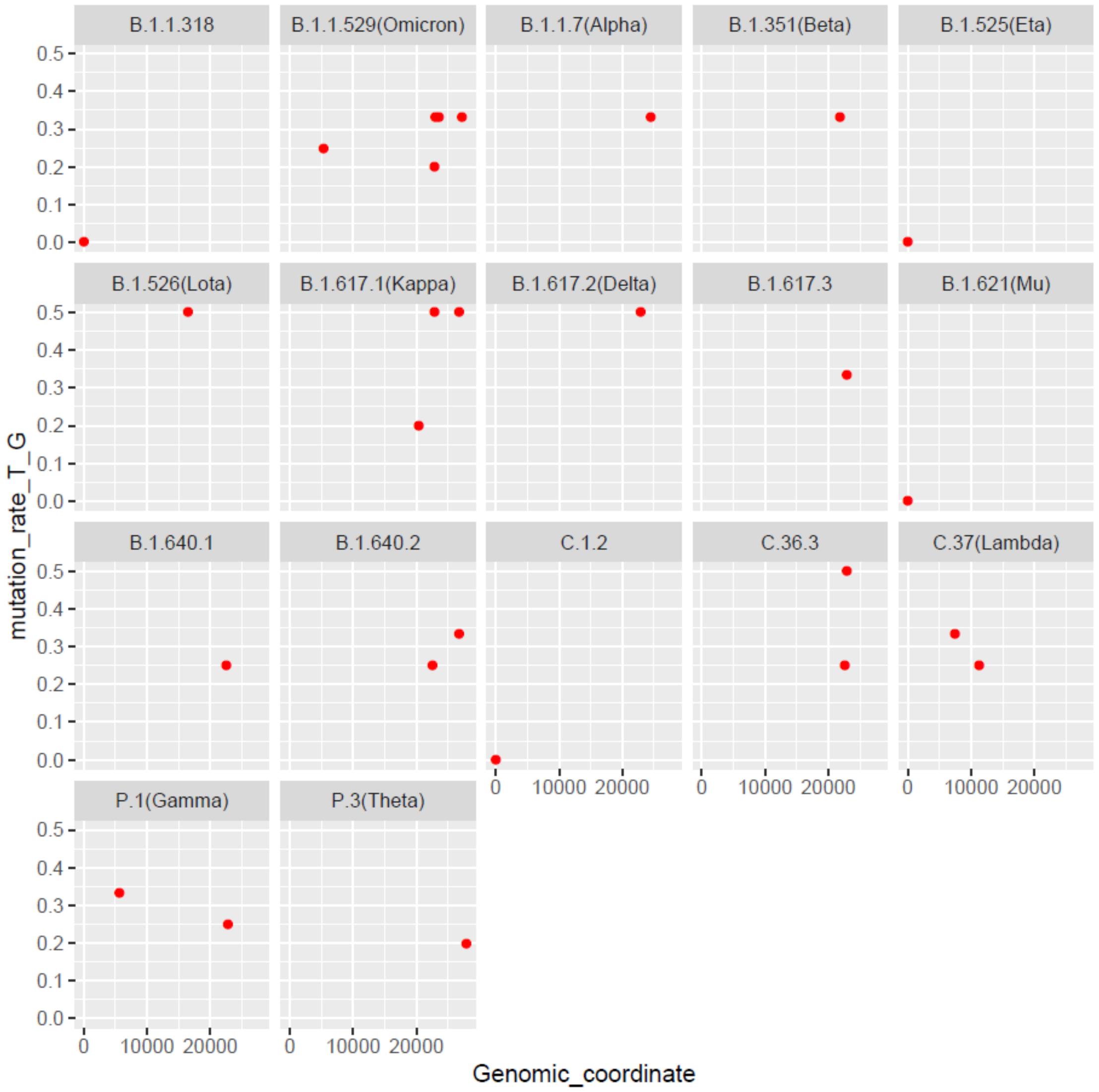

**Supplementary Figure 17. The richness distribution of the mutation rate of G->C across the whole SARS-CoV-2 genome in different lineages.** The same as above Supplementary Figure 12.

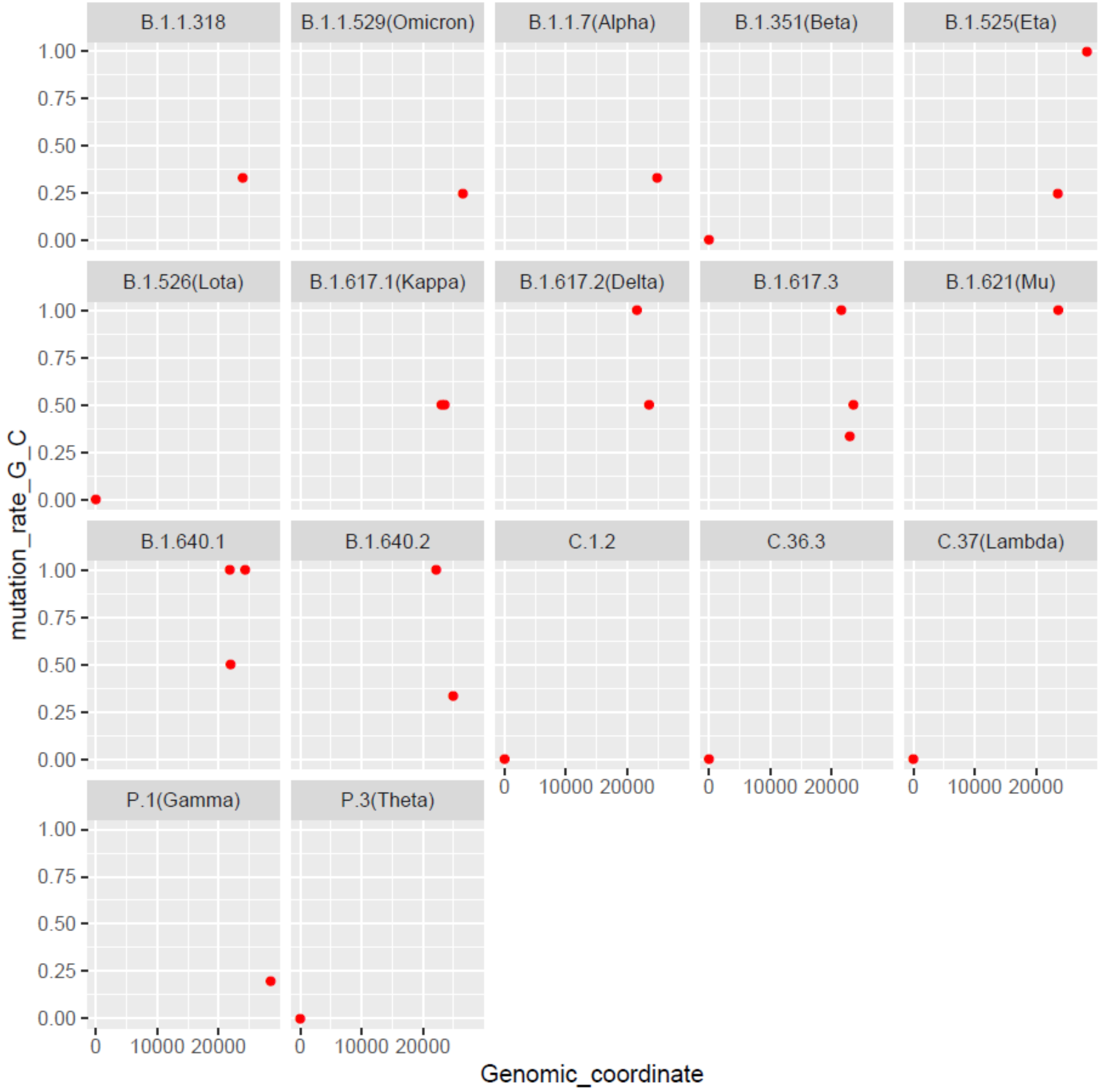

**Supplementary Figure 18. Smoothed distribution of new cases per million cases of SARS-CoV-2 in different severely affected areas, such as Africa, Asia, Europe, North America, Oceania, and South America, along the timeline.**

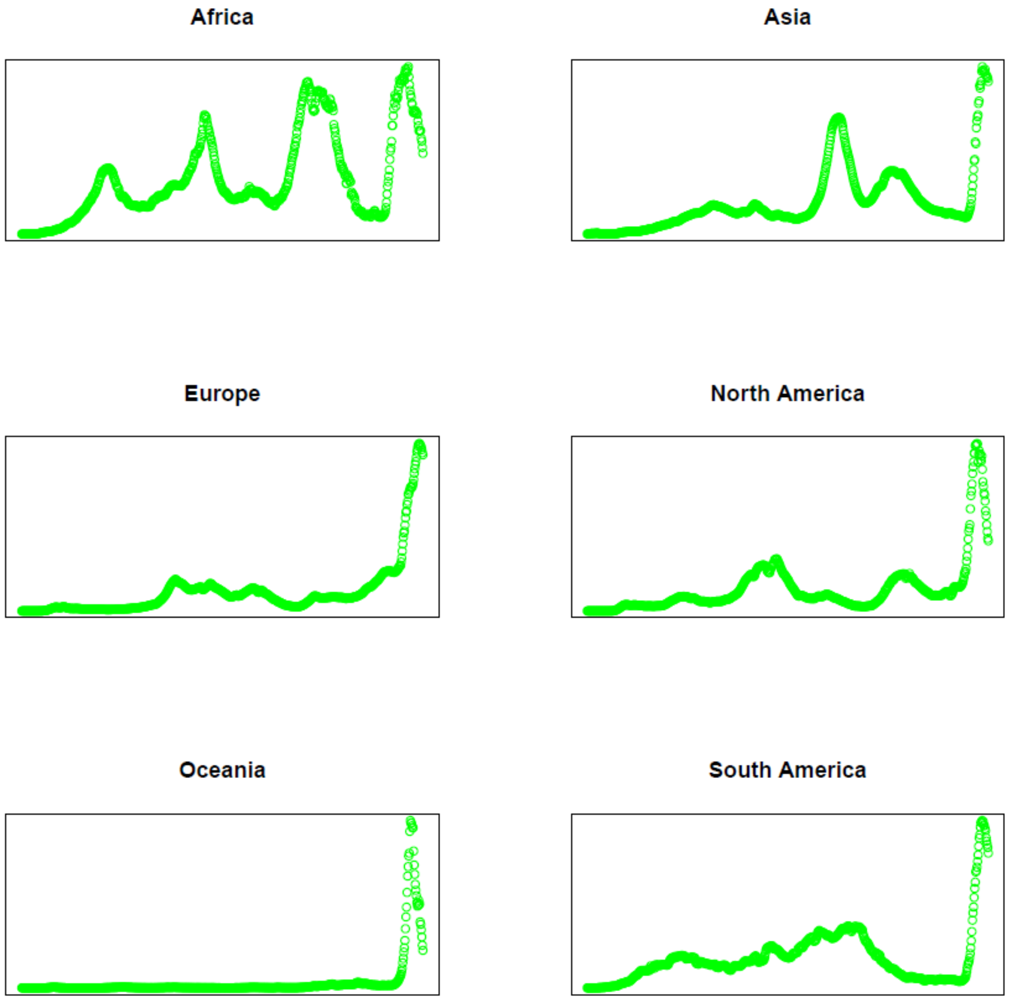

**Supplementary Figure 19. Smoothed distribution of new cases per million cases of SARS-CoV-2 in Africa along the timeline.** Red boxes represent no continued outbreaks excluded in our analysis.

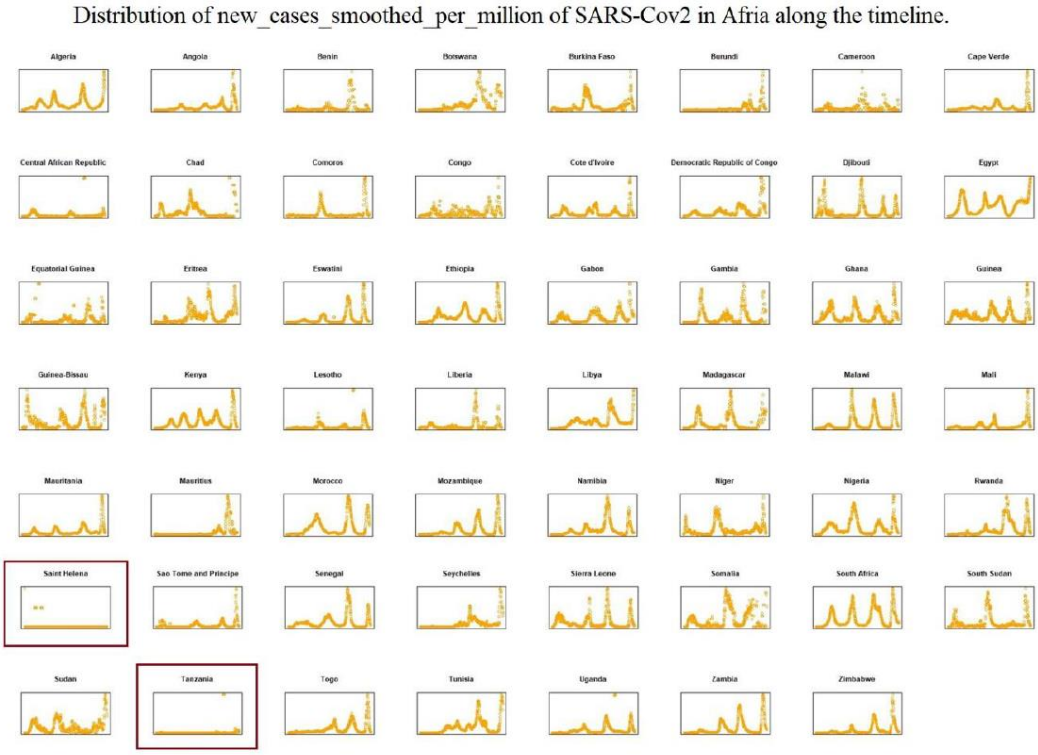

**Supplementary Figure 20. Smoothed distribution of new cases per million cases of SARS-CoV-2 in Asia along the timeline.**

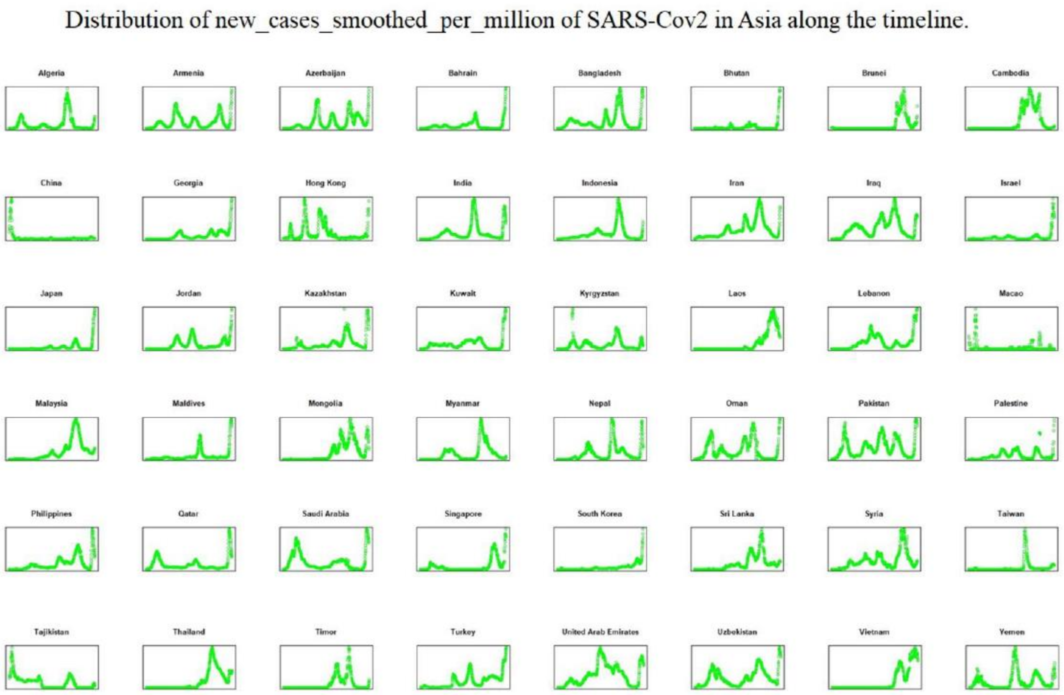

**Supplementary Figure 21. Smoothed distribution of new cases per million cases of SARS-CoV-2 in South America along the timeline.** Red boxes represent no continued outbreaks excluded in our analysis.

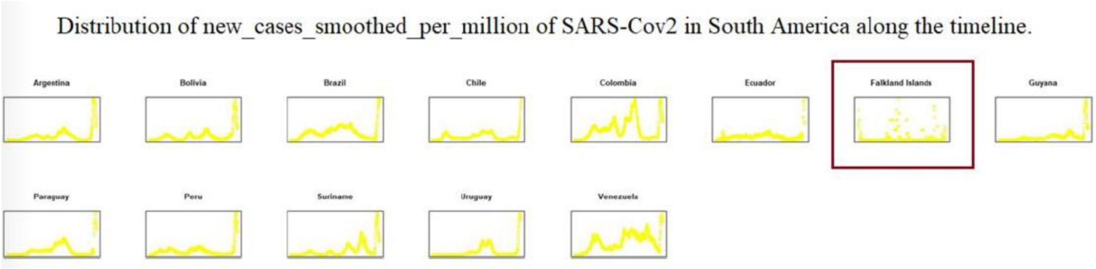

**Supplementary Figure 22. Smoothed distribution of new cases per million cases of SARS-CoV-2 in Europe along the timeline.** Red boxes represent no continued outbreaks excluded in our analysis.

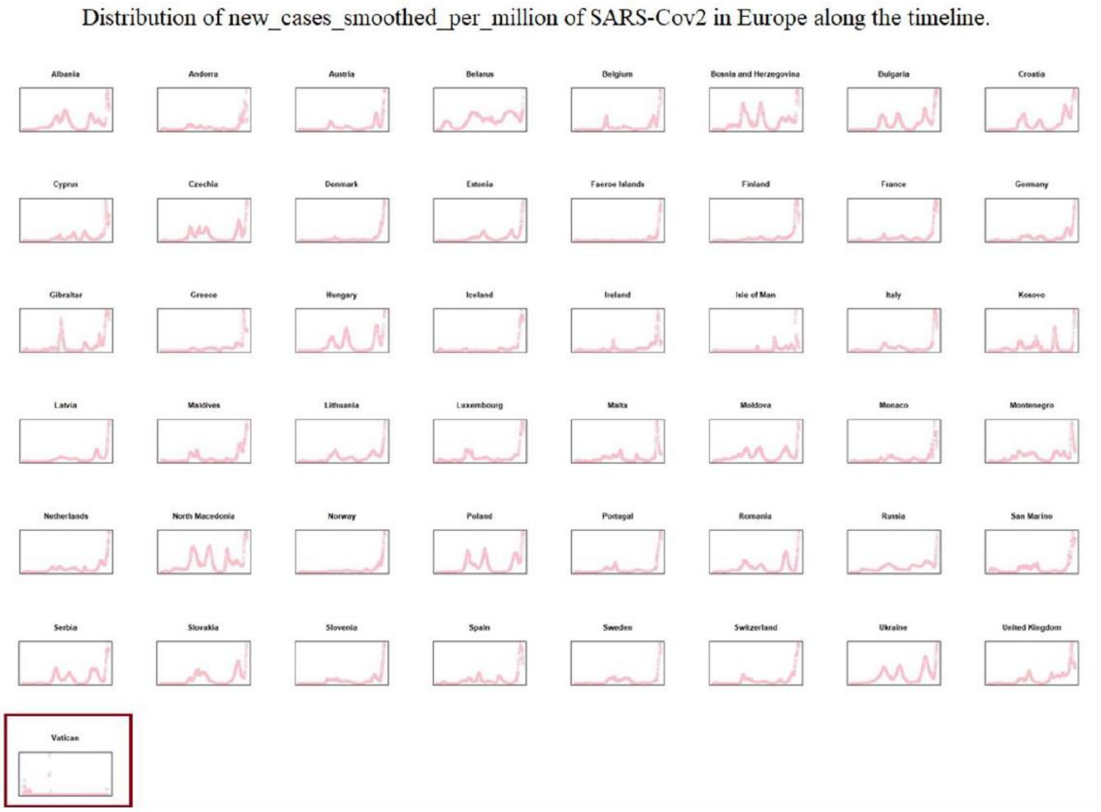

**Supplementary Figure 23. Smoothed distribution of new cases per million cases of SARS-CoV-2 in North America along the timeline.** Red boxes represent no continued outbreaks excluded in our analysis.

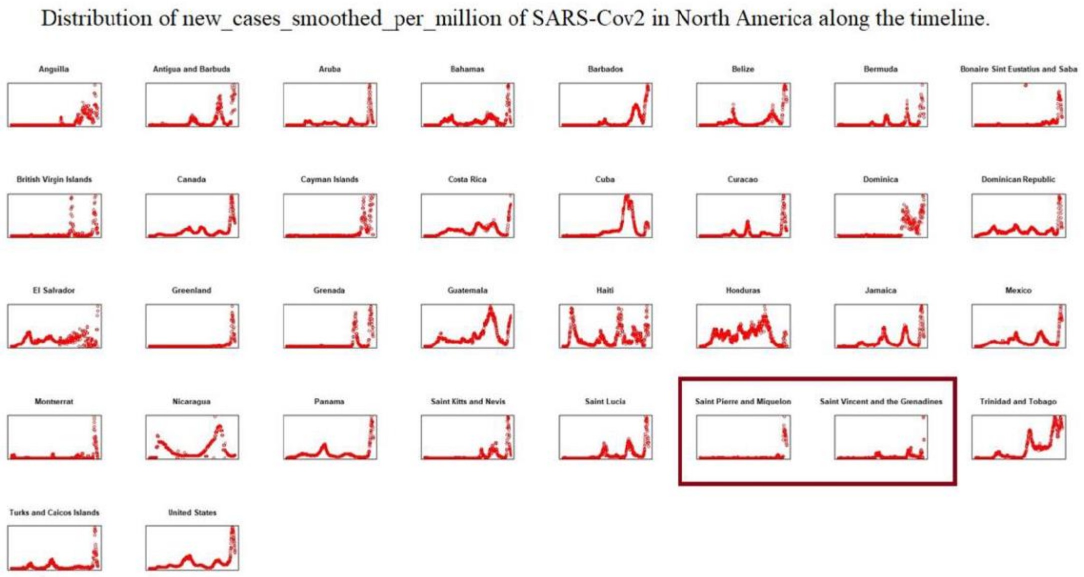

**Supplementary Figure 24. Smoothed distribution of new cases per million cases of SARS-CoV-2 in Oceania along the timeline.** Red boxes represent no continued outbreaks excluded in our analysis.

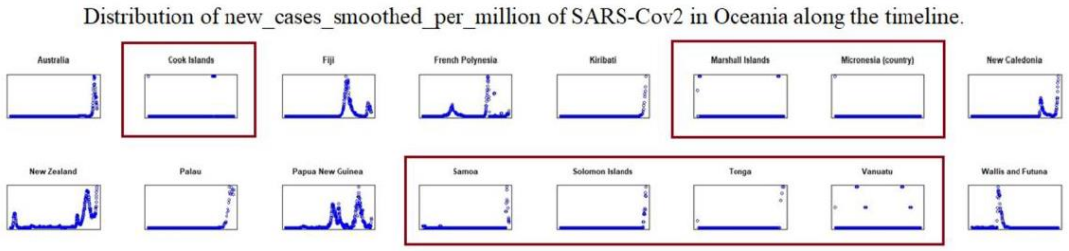

**Supplementary Figure 25. Frequency distribution of different types for substitutions 1-20.** Different types of amino acid substitutes are represented by different colours, such as A (Gla), C (Cys), D (Asp), E (Glu), F (Phe), G (Gly), H (His), I (Ile), K (Lys), L (Leu), M (Met), N (Asn), P (Pro), Q (Gln), R (Arg), S (Ser), Stop, T (Thr), V (Val), W (Trp), Y (Tyr).

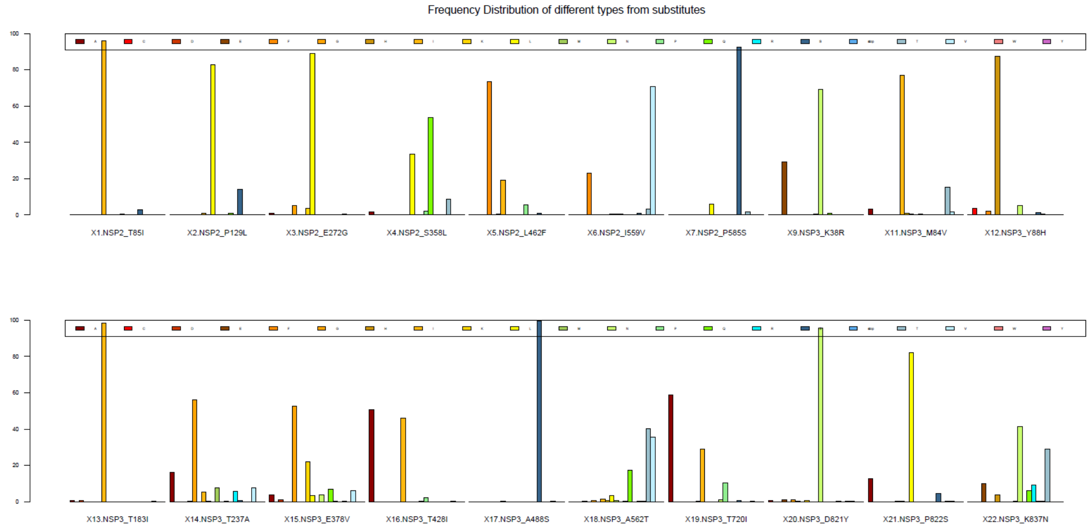

**Supplementary Figure 26. Frequency distribution of different types for substitutions 21-40.** Different types of amino acid substitutes are represented by different colours, such as A (Gla), C (Cys), D (Asp), E (Glu), F (Phe), G (Gly), H (His), I (Ile), K (Lys), L (Leu), M (Met), N (Asn), P (Pro), Q (Gln), R (Arg), S (Ser), Stop, T (Thr), V (Val), W (Trp), Y (Tyr).

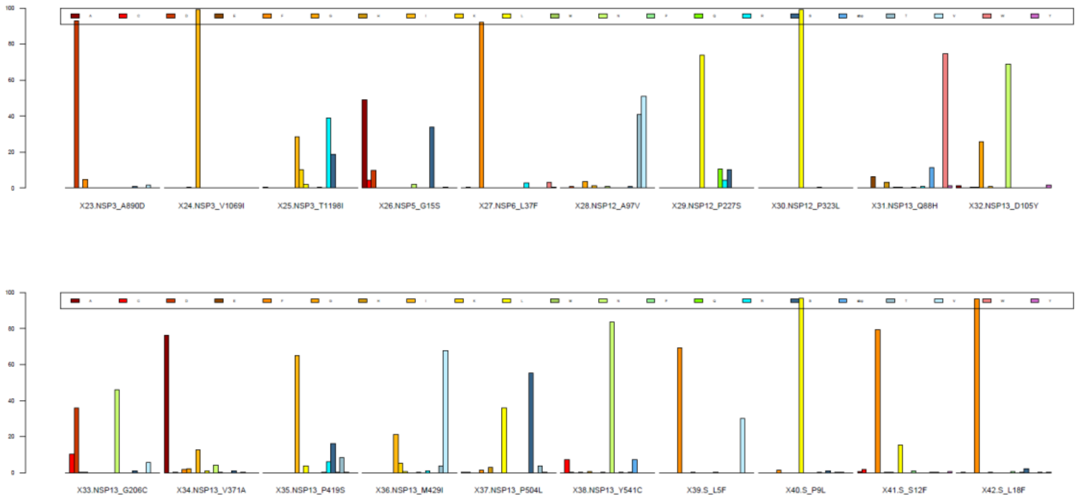

**Supplementary Figure 27. Frequency distribution of different types for substitutions 41-60.** Different types of amino acid substitutes are represented by different colours, such as A (Gla), C (Cys), D (Asp), E (Glu), F (Phe), G (Gly), H (His), I (Ile), K (Lys), L (Leu), M (Met), N (Asn), P (Pro), Q (Gln), R (Arg), S (Ser), Stop, T (Thr), V (Val), W (Trp), Y (Tyr).

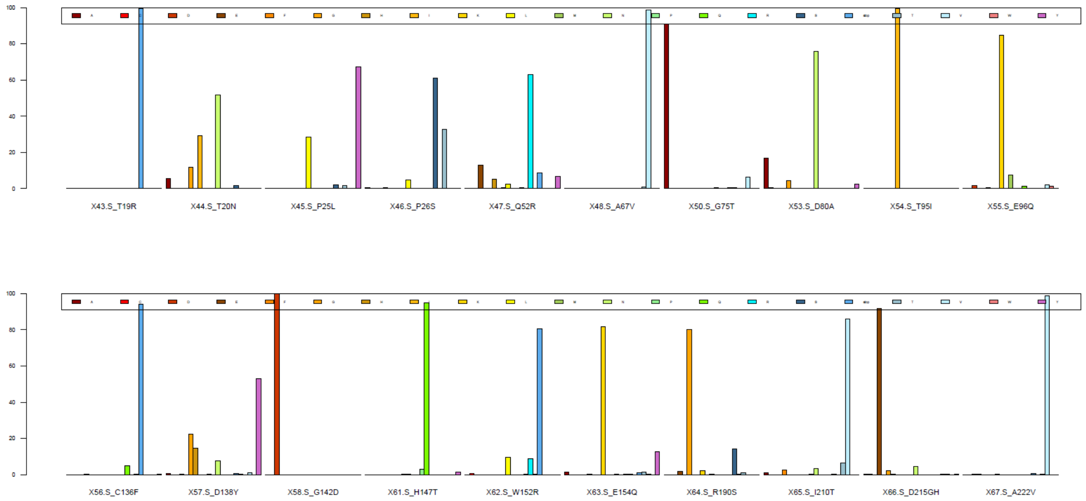

**Supplementary Figure 28. Frequency distribution of different types for substitutions 61-80.** Different types of amino acid substitutes are represented by different colours, such as A (Gla), C (Cys), D (Asp), E (Glu), F (Phe), G (Gly), H (His), I (Ile), K (Lys), L (Leu), M (Met), N (Asn), P (Pro), Q (Gln), R (Arg), S (Ser), Stop, T (Thr), V (Val), W (Trp), Y (Tyr).

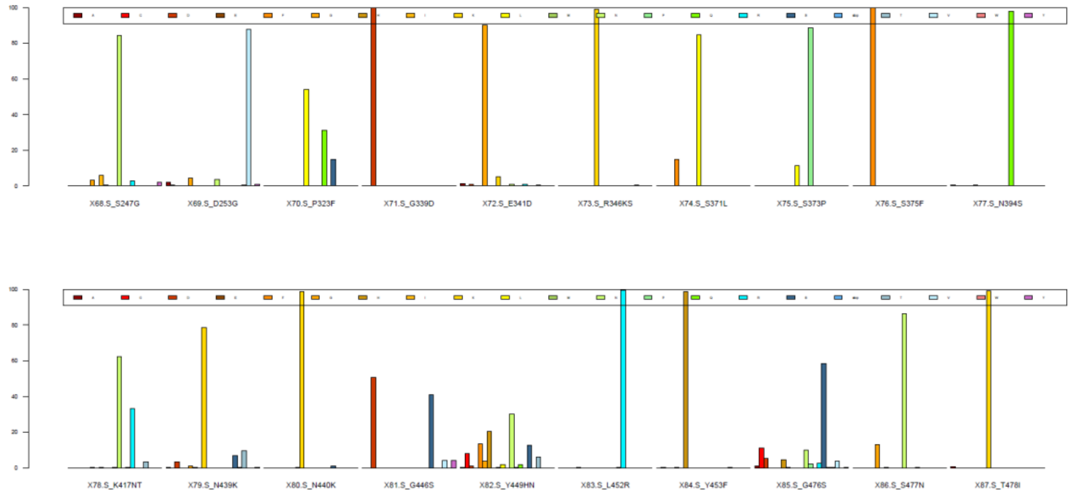

**Supplementary Figure 29 Frequency distribution of different types for substitutions 81-100.** Different types of amino acid substitutes are represented by different colours, such as A (Gla), C (Cys), D (Asp), E (Glu), F (Phe), G (Gly), H (His), I (Ile), K (Lys), L (Leu), M (Met), N (Asn), P (Pro), Q (Gln), R (Arg), S (Ser), Stop, T (Thr), V (Val), W (Trp), Y (Tyr).

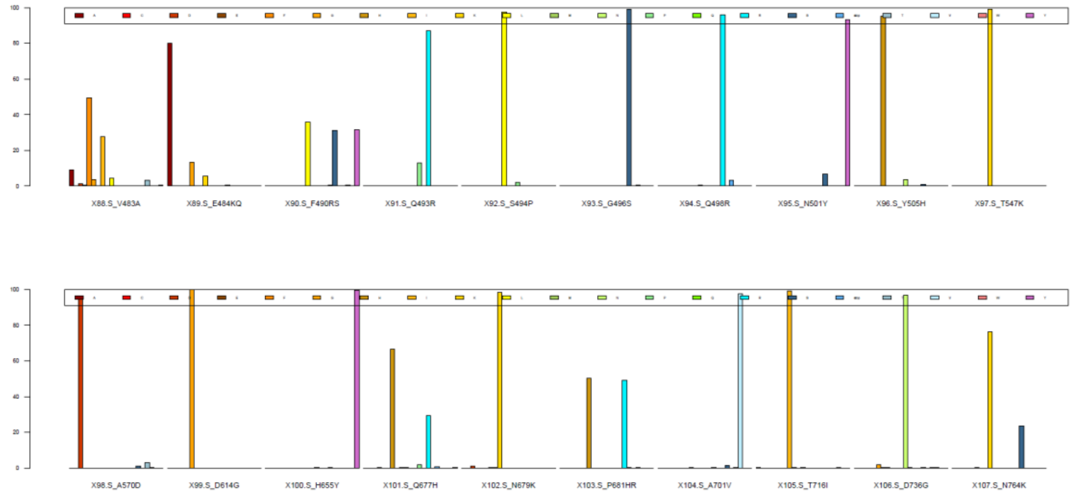

**Supplementary Figure 30. Frequency distribution of different types for substitutions 101-120.** Different types of amino acid substitutes are represented by different colours, such as A (Gla), C (Cys), D (Asp), E (Glu), F (Phe), G (Gly), H (His), I (Ile), K (Lys), L (Leu), M (Met), N (Asn), P (Pro), Q (Gln), R (Arg), S (Ser), Stop, T (Thr), V (Val), W (Trp), Y (Tyr).

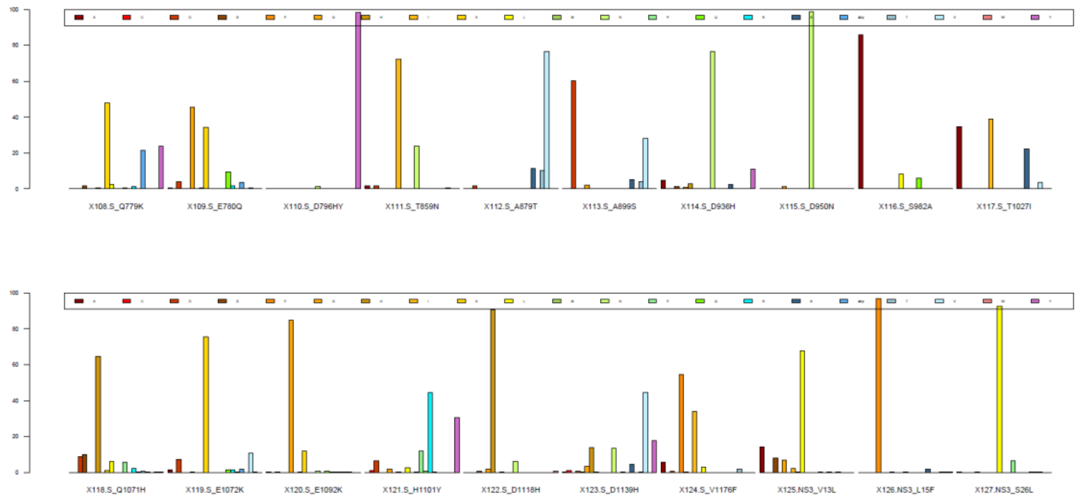

**Supplementary Figure 31. Frequency distribution of different types for substitutions 121-140.** Different types of amino acid substitutes are represented by different colours, such as A (Gla), C (Cys), D (Asp), E (Glu), F (Phe), G (Gly), H (His), I (Ile), K (Lys), L (Leu), M (Met), N (Asn), P (Pro), Q (Gln), R (Arg), S (Ser), Stop, T (Thr), V (Val), W (Trp), Y (Tyr).

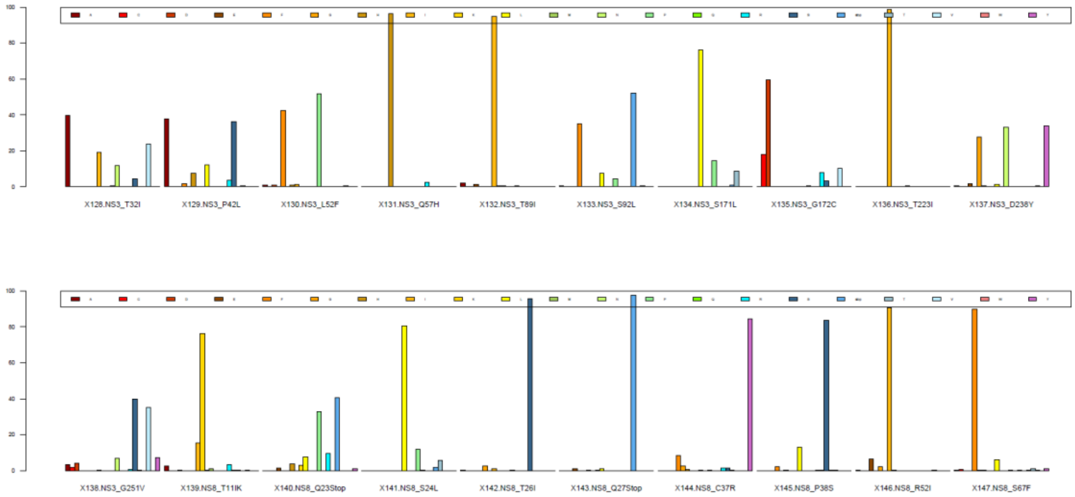

**Supplementary Figure 32. Frequency distribution of different types for substitutions 141-158.** Different types of amino acid substitutes are represented by different colours, such as A (Gla), C (Cys), D (Asp), E (Glu), F (Phe), G (Gly), H (His), I (Ile), K (Lys), L (Leu), M (Met), N (Asn), P (Pro), Q (Gln), R (Arg), S (Ser), Stop, T (Thr), V (Val), W (Trp), Y (Tyr).

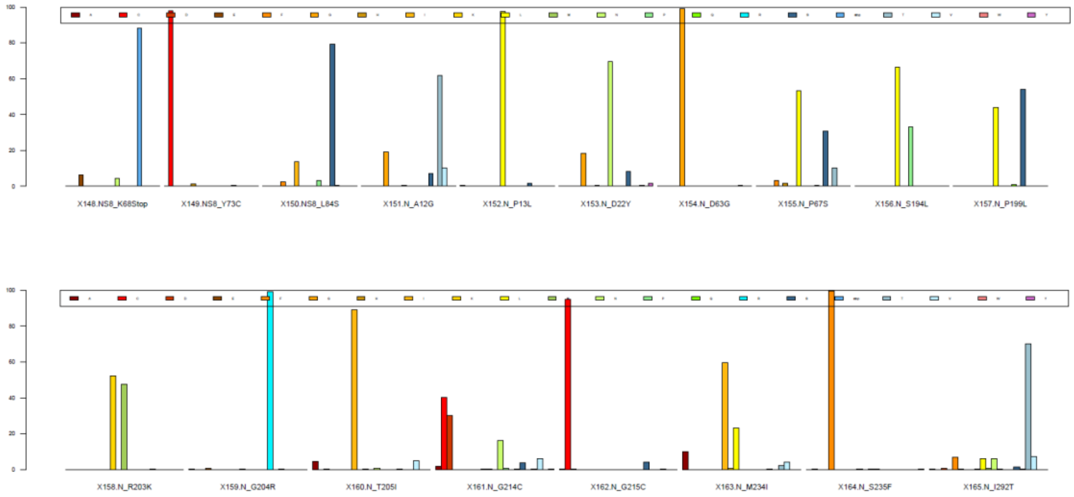

**Supplementary Table 1. Comprehensive information on *Coronaviridae*, especially different lineages of SARS-CoV-2.** Sheet “Plague” contains data on the outbreak of epidemic disease in human history. Sheet “Cov” contains the taxonomy of *Coronaviridae* according to the NCBI. Sheet “branch” contains the studied groups related to the outgroup on the evolutionary branch of *Coronaviridae*. Sheet “host” contains the basic information of hosts of *Coronaviridae*. Sheet “AA substitutes” contains specific amino acid substitutions with different mutation types in different lineages of SARS-CoV-2. Sheet “Codon Usage” contains the codon numbers of different lineages of SARS-CoV-2. Sheet “RSCU” contains RSCU values for different lineages of SARS-CoV-2. Sheet “lineages” contains the global distribution of lineages of SARS-CoV-2.

**Supplementary Table 2. The comprehensive information of true sets.** Sheets include the distribution of a total of 144 amino acid substitutions in different lineages, new cases per million from different countries/regions on different continents, and the statistics of different mutation types of amino acid substitutes.

**Supplementary Table 3. Comprehensive information of ZHU prediction.** Sheets include 75 training sets and 43 validation sets, the information on the input data, GLM coefficient estimates and AIC values, 171 significant substitutions, prediction results for the training sets, GLM and reordered processing data, the ZHU prediction model, prediction results for the validation sets, and performance parameters.
